# The Pandemic’s Echo: A Population-Based Assessment of MMR Rates and Associated Factors in the Wake of COVID-19

**DOI:** 10.1101/2025.02.13.25322239

**Authors:** Ensheng Dong, Samee Saiyed, Andreas Nearchou, Yamato Okura, Lauren M. Gardner

## Abstract

**Objectives:** We generate a high-resolution data set on MMR vaccination coverage in the U.S. before and after the COVID-19 pandemic. We use this data to conduct a spatiotemporal analysis of vaccination patterns and uncover the associations between MMR vaccination rates and key factors such as socioeconomic conditions, COVID-19 impact, vaccine policy and other health-related variables.

**Methods:** We collect county-level 2-dose MMR vaccination rates for kindergarten children from 2017 – 2024 for 2,266 U.S. counties, compare coverage patterns and trends before and after the COVID-19 pandemic, and implement multivariable weighted logistic regression models to identify factors associated with low and declining MMR vaccination rates in U.S. counties.

**Results:** We reveal a nationwide decline in MMR rates following the COVID-19 pandemic, with declines observed in 1,688 of the counties evaluated. We find state-level non-medical exemption (NME) policies to be the strongest associated factor with low and declining MMR rates at the county level post-pandemic. We also identify a positive association between county-level MMR rates and uptake of other vaccine types, the minority proportion in a county and Republican-aligned counties. In contrast, county-level MMR rates are negatively associated with post-secondary education rates. In addition to NMEs, MMR rate declines are positively associated with the proportion of rural population and a higher religious diversity at the county level, and negatively associated with household income and the proportion of Latinx people.

**Conclusions:** The significant association between NMEs and low and declining MMR rates suggest these state-level policies are harmful and lead to reduced vaccination coverage post-pandemic. The positive associations among vaccine types suggest spillover effects in vaccine seeking behavior. The association between MMR rates and Republican-aligned counties contrast COVID-19 vaccination patterns, highlighting the complex nature of political polarization on vaccine-related behavior.

## Introduction

It is widely accepted in the scientific communities that childhood vaccination is an effective and efficient public health intervention, which has substantially reduced the incidence of vaccine-preventable diseases and has saved countless children’s lives worldwide.^1^ In the United States (U.S.), the Advisory Committee on Vaccination Practices (ACIP) recommends a series of vaccinations for children and adolescents to protect against various infectious diseases.^2^ Researchers estimated that the childhood vaccination program prevented over 24 million total cases of vaccine-preventable diseases for the U.S. population in 2019 alone.^3^ Nonetheless, in the U.S. there is evidence of a national-level decline in the measles-mumps-rubella (MMR) vaccination rate among children between 2019 and 2024 (Figure 1).^4^ Furthermore, the Centers for Disease Control and Prevention (CDC) has reported a resurgence of measles outbreaks,^4^ which has typically served as a sentinel indicator of childhood vaccine interruptions.^5^ As of March 27, 2025 there are 483 confirmed measles cases reported across 20 jurisdictions (already surpassing the total number of cases in 2024), with large outbreaks ongoing in Texas and New Mexico, and the vast majority of the cases occurring in unvaccinated children.^4^

**Figure 1.**
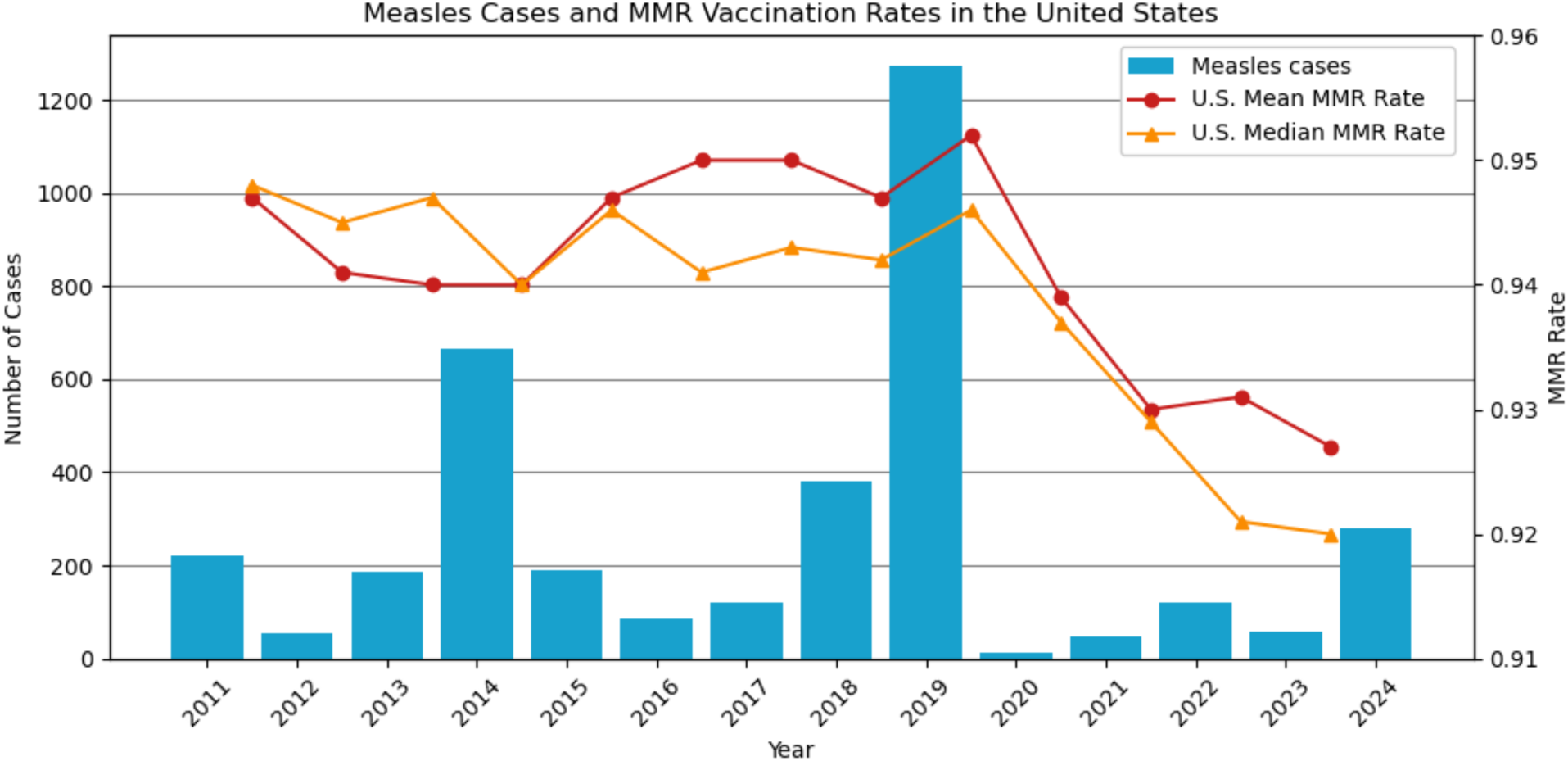
Reported measles cases from 2011 to 2024 and national MMR vaccination rates in the United States from school year 2011-12 to 2023-24. (Source: CDC,^4^ as of December 8, 2024)

Various factors have been shown to impact childhood vaccination rates in the U.S, including parental vaccine hesitancy, socioeconomic disparities, political affiliation, policy differences across states, and online misinformation.^5–9^ Beyond the more traditional determinants of vaccination choice, the COVID-19 pandemic further exacerbated the decline in routine childhood vaccination coverage, potentially due to the disruption of healthcare services and the diversion of public health resources.^10,11^ Furthermore, the widespread vaccine hesitancy observed during the COVID-19 pandemic in the U.S. - characterized by significant regional disparities in vaccination uptake and strong influences of political and social factors – has raised concerns about potential spillover effects on routine childhood vaccination acceptance and coverage.^12–16^ Several studies have examined childhood vaccination during the pandemic period, reporting decreases in pediatric vaccine ordering, childhood vaccination coverage, and up-to-date vaccination status,^10,17,18^ however, the first two studies only focus on the acute early phase of the COVID-19 pandemic while the latter only examines specific U.S. regions. A survey-based study by Zhou et al conducted in 2023-24 revealed associations between parental characteristics – such as socio-demographics, political affiliation, and COVID-19 vaccination status and MMR vaccination choice, however these relationships have yet to be evaluated at a population level, within a comprehensive nationwide study. Additionally, previous research has raised significant concerns about the impact of state-level vaccination policies on declining MMR vaccination coverage.^19,20^ In particular, non-medical exemptions (NMEs), which are classified as religious or philosophical exemptions from vaccination requirements, have been strongly associated with declining MMR coverage and identified as a contributing factor to measles outbreaks in the U.S.^8,21–23^

While the national and state level declines in MMR coverage over the last five years are well documented,^4^ outbreaks occur within communities, which may differ in their vaccination coverage substantially, even within a state. However, there are no comprehensive datasets or studies evaluating nationwide MMR coverage at a county-level in the post-COVID era. In this study we build on the existing literature, addressing previous limitations in scale and resolution to provide a more comprehensive understanding of the geographical heterogeneity in MMR vaccine uptake and its underlying determinants. Specifically, the main contributions of our study are as follows:

1. We conducted a comprehensive data collection effort to generate a standardized dataset with annual county-level 2-dose MMR vaccination rates from 2017 to 2024 for all states in the U.S. where this information is available.
2. We evaluated spatiotemporal trends in MMR coverage to reveal the U.S. counties with low MMR vaccination rates or significant declines during the pandemic period.
3. We employed multivariable weighted logistic regression models to reveal the factors associated with low and declining MMR vaccination rates post-pandemic.

This study is unique in its population-based approach, high spatial resolution (county-level) and inclusion of a comprehensive set of explanatory factors, enabling a deeper understanding of nationwide childhood vaccination rates before and after the COVID-19 pandemic. It offers critical insights for policymakers to address local-level declines in vaccination rates, inform policies that enhance the resilience of the childhood vaccination program, and implement effective strategies to help children catch up on missed vaccinations where necessary.

## Methods

In this section, we describe our extensive data collection efforts to compile county-level MMR vaccination data from 2017 to 2024, as well as county-level data on demographics, socioeconomics, political affiliation, (state-level) vaccine policy, COVID-19 metrics and other health-related factors. We describe our methodology for identifying counties with significant changes in MMR vaccination rates and outline the multivariable logistic regression models implemented to reveal determinants of low and declining MMR vaccination rates during the study period.

### Data collection and standardization of county-level MMR vaccination rates for the U.S

The primary variable of interest in this study is county-level MMR vaccination rates for school aged children, specifically the 2-dose MMR vaccination rates for kindergarteners. This metric represents completion of the full MMR vaccination series, where the first dose is typically administered near age one and the second near age five, presumably right before entering kindergarten. Where available, we collected this metric (or the best available proxy) from each state’s Department of Health website for each of the school years 2017-18 to 2023-24. Our study includes 37 states covering 2,266 counties. To ensure consistency across states, we developed a standardized data collection procedure that accounted for the various discrepancies in reporting. A detailed description of the data collection process and standardization is documented in Supplementary Section A and Table S1. The final dataset enables a high-resolution analysis of MMR vaccine coverage patterns across the U.S.

### Data collection for modeling determinants of MMR vaccination rates

In addition to collecting and analyzing MMR rates and trends, we seek to determine the factors associated with low and declining MMR vaccination rates in U.S. counties. We therefore collect a series of county- and state-level variables previously found to be associated with MMR rates in the literature.^7,9,13,24^ The set of variables is classified into four main categories, as listed in Table S2: 1) demographic and socioeconomic factors, 2) political affiliation, 3) health-related factors including vaccine uptake for other vaccine types, COVID-19 health outcomes, and a proxy for healthcare accessibility, and 4) vaccine policy, specifically state-level non-medical exemption (NME) policies. A detailed description of each variable is provided in Supplementary Section A.

### Spatial-temporal trend analysis of MMR vaccination rates

We use our comprehensive county-level MMR coverage dataset to identify trends and significant changes in MMR vaccination rates at the county-level, specifically those counties with low coverage, as well as counties experiencing significant declines in MMR vaccination rates during the pandemic.

### Statistical analysis to reveal factors associated with low and declining MMR vaccination rates in the U.S

To improve our understanding of the factors associated with low MMR vaccination rates (i.e., rates below the critical 95% threshold), and how the factors might differ pre-and post-pandemic in U.S. counties, we developed two multivariable weighted logistic regression models: 1) MMR Pre-Pandemic Model, and 2) MMR Post-Pandemic Model. Additionally, we developed a third model, 3) MMR Declining Model, to reveal the factors associated with declines in MMR rates between the pre- and post-pandemic period. The objective of each model and their baseline reference groups are specified below. The total number of counties included in each model is 2266.

1. **MMR Pre-Pandemic Model:** Identifies factors associated with those counties reporting low MMR vaccination rates (i.e., less than or equal to 95%) before the pandemic. Counties with MMR rates equal to or below the 95% coverage pre-pandemic serve as the baseline reference group, comprising 1057 counties.
2. **MMR Post-Pandemic Model:** Identifies factors associated with those counties reporting low MMR vaccination rates (i.e., less than or equal to 95%) after the pandemic. Counties with MMR rates equal to or below the 95% coverage post-pandemic serve as the baseline reference group, comprising 1515 counties.
3. **MMR Declining Model:** Identifies factors associated with those counties that experienced substantial declines in MMR vaccination rates between the pre-and post-pandemic periods, specifically defined as any decrease equal to or greater than the national county-level average of 2.36% during this period. Counties with an MMR rate decline of at least 2.36% serve as the baseline reference group, comprising 977 counties.

We selected 95% as the threshold to determine the baseline reference groups for the MMR Pre- and Post-Pandemic Models because it is widely used in academic publications and government policies as the minimum MMR vaccine target for herd immunity.^25^ To assess the robustness of our findings to this choice we conducted a sensitivity analysis for this threshold, which is presented in Supplementary Section C. Similarly, we conduct additional sensitivity analysis on the chosen rate (2.36%) used to define the baseline group for the MMR Declining Model, which is also presented in Supplementary Section C. Figure S3 visually depicts the spatial distribution of the baseline reference groups corresponding to the selected thresholds for each model. Lastly, in addition to modeling the pre-and post-pandemic periods independently we also implemented a unified pre- and post-pandemic logistic regression model, which is presented in Supplementary Section B and results shown in Supplementary Section C. The results from the joint model are consistent with the period-specific models, however for purposes of interpretability, we focus our results and discussion on period-specific models.

To mitigate noise in reporting and to capture robust representations of pre-and post-pandemic MMR rates, we use averages of annual MMR vaccination rates over specified time windows to define the static pre-and post-pandemic MMR rates used in the model. Specifically for each county, the pre-pandemic MMR vaccination rate is the average of the rates for the 2017-18, 2018-19, and 2019-20 school years, while the post-pandemic MMR rate is the average of the rates for the 2022-23 and 2023-24 school years for each county (when the data is available). A comprehensive overview of each model, input data, and relevant limitations can be found in Supplementary Section B. All three models incorporate a wide range of explanatory variables (summarized in Table S2, with the correlation matrix shown in Figure S4).

## Results

### County-level MMR vaccination rates before and after the COVID-19 pandemic

Figure 2A and 2B illustrates the county-level MMR vaccination rates before and after the pandemic, respectively, while Figure 2C illustrates the distribution of MMR vaccination rates for all counties in each reporting state. Our findings reveal a trending nationwide decline in MMR vaccination rates in the U.S. over the course of the COVID-19 pandemic, with significant heterogeneity within and across states. The county-level average MMR rate fell from 93.89% pre-pandemic to 91.53% post-pandemic – an average decline of 2.36%. Specifically, 1,688 counties out of 2,266 studied counties report a decline in coverage after the pandemic, with 977 counties reporting declines equal to or greater than the national average of 2.36%. Further, 458 additional counties fell below the 95% threshold post-pandemic compared to the pre-pandemic period, increasing the total from 1,057 pre-pandemic to 1,515 post-pandemic. Notably, only four of the 37 states in our study, namely Maine, New York, Connecticut, and New Mexico, reported an increase in the median county-level MMR rate (Figure 2B). These county-level results reinforce the trends from state and national level reporting by the CDC, with 43 states out of 49 states reporting drops in rates (Montana has no post-pandemic data).

**Figure 2.**
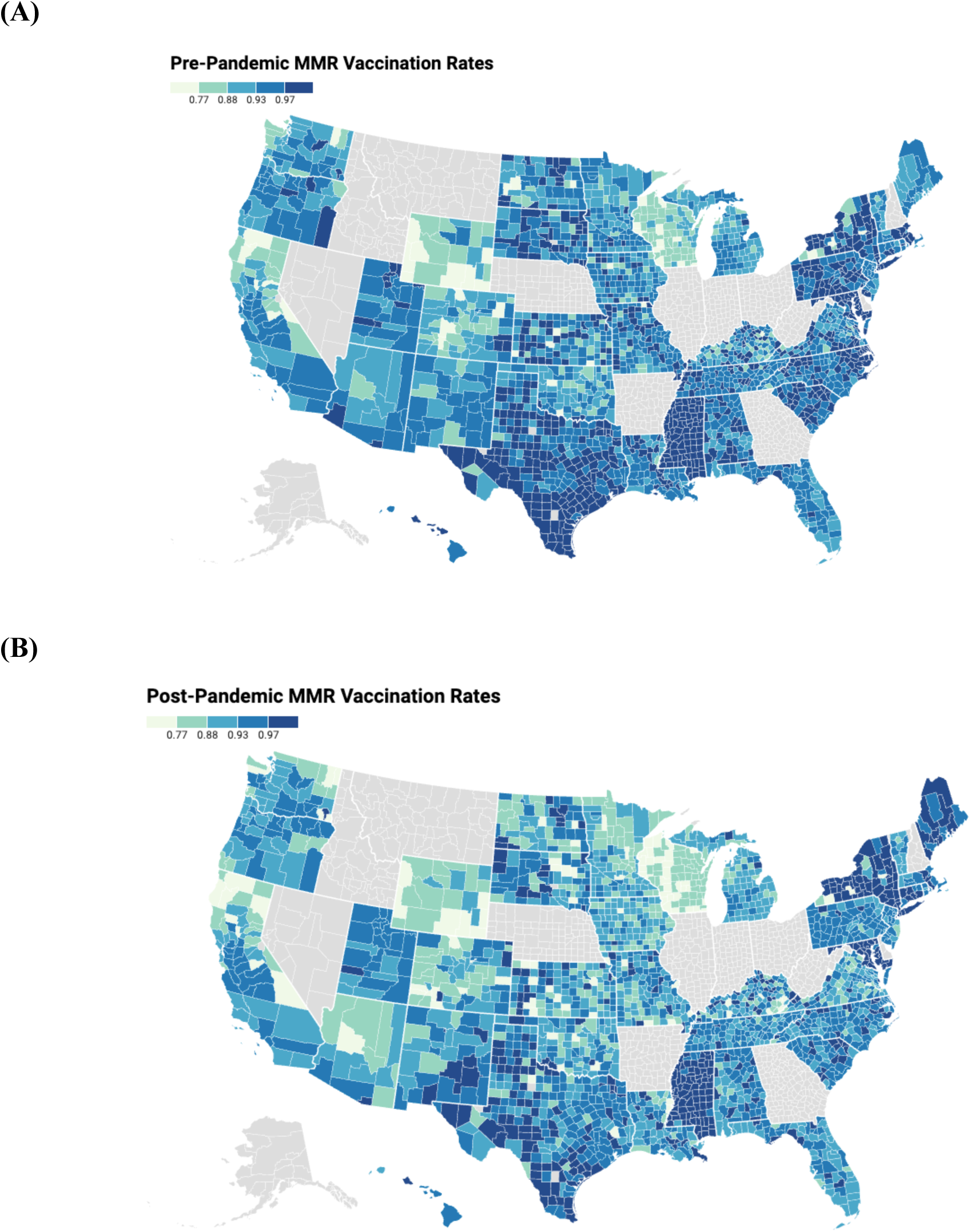
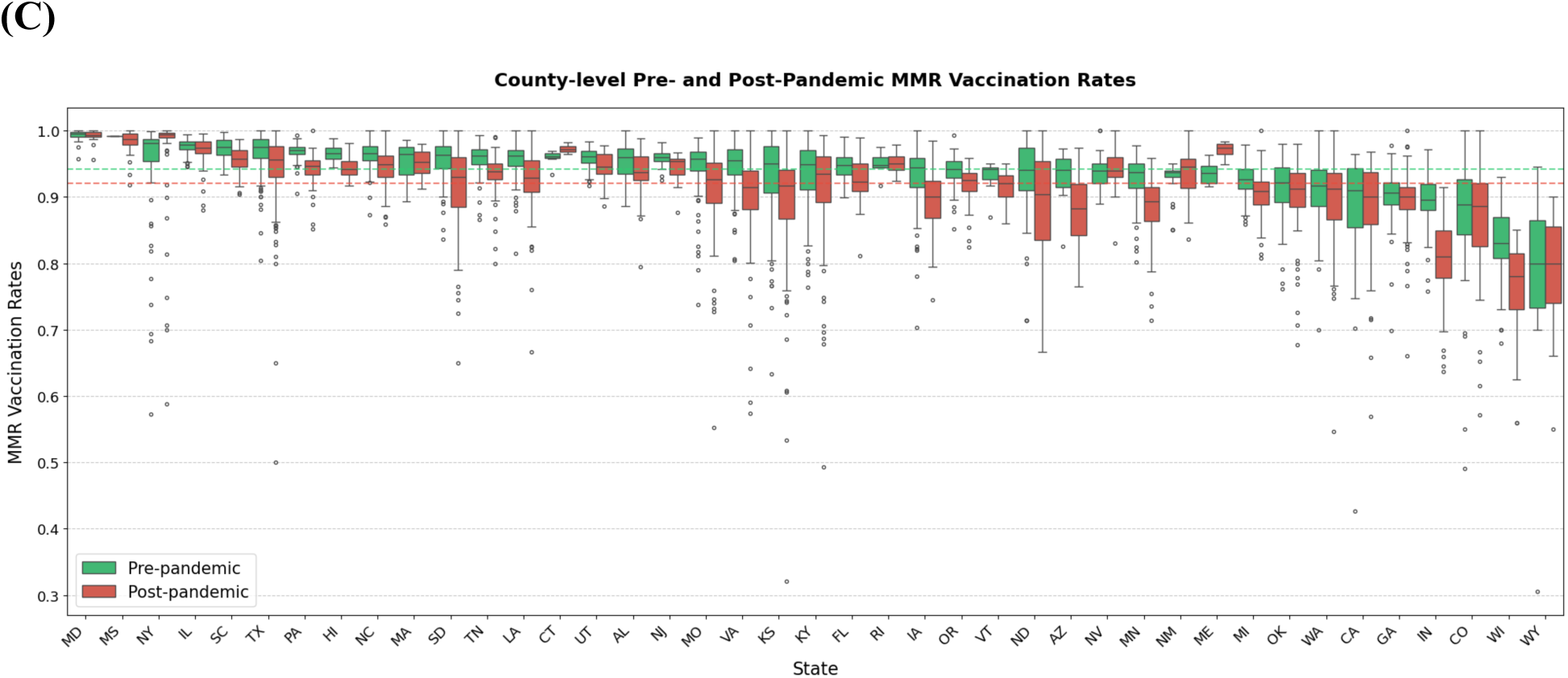
Maps plotting the county-level MMR vaccination rates before (Panel A) and after (Panel B) the COVID-19 pandemic. Counties in grey are those with missing MMR data. Please refer to Supplementary A for a detailed description of the data collection procedure. Maps are created using Datawrapper.^42^ Panel (C) presents a boxplot showing the county-level MMR vaccination coverages for each state before (green) and after (orange) the pandemic. Each boxplot shows the distribution of the MMR vaccination rates for all counties in a state. Boxplots are ranked from the states with the highest to the lowest median MMR vaccination rates. The national average (from all 50 states and D.C.) of 2-dose kindergarten MMR vaccination rate is 95% (green-line) and 93% (orange-line) before and after the pandemic respectively,^4^ delineated by two horizonal dashed lines.

### Factors associated with low MMR vaccination rates before and after the pandemic

The modeled associations from the MMR Pre-Pandemic Model and MMR Post-Pandemic Model are represented as odds ratios in Figure 3 and Table S5. The MMR Pre-Pandemic Model indicates that counties with higher postsecondary education rates are more likely to have MMR vaccination rates below 95%. In contrast, higher percentages of Black and Latinx populations, a higher relative median household income, a greater percentage of flu-vaccinated fee-for-service Medicare enrollees (FFS flu vaccinated percentage), a higher healthcare accessibility (measured by hospitals and pharmacies per 1,000 people), and a larger percentage of voters supporting Donald Trump in the 2020 presidential election are associated with counties having MMR vaccination rates above 95%.

**Figure 3.**
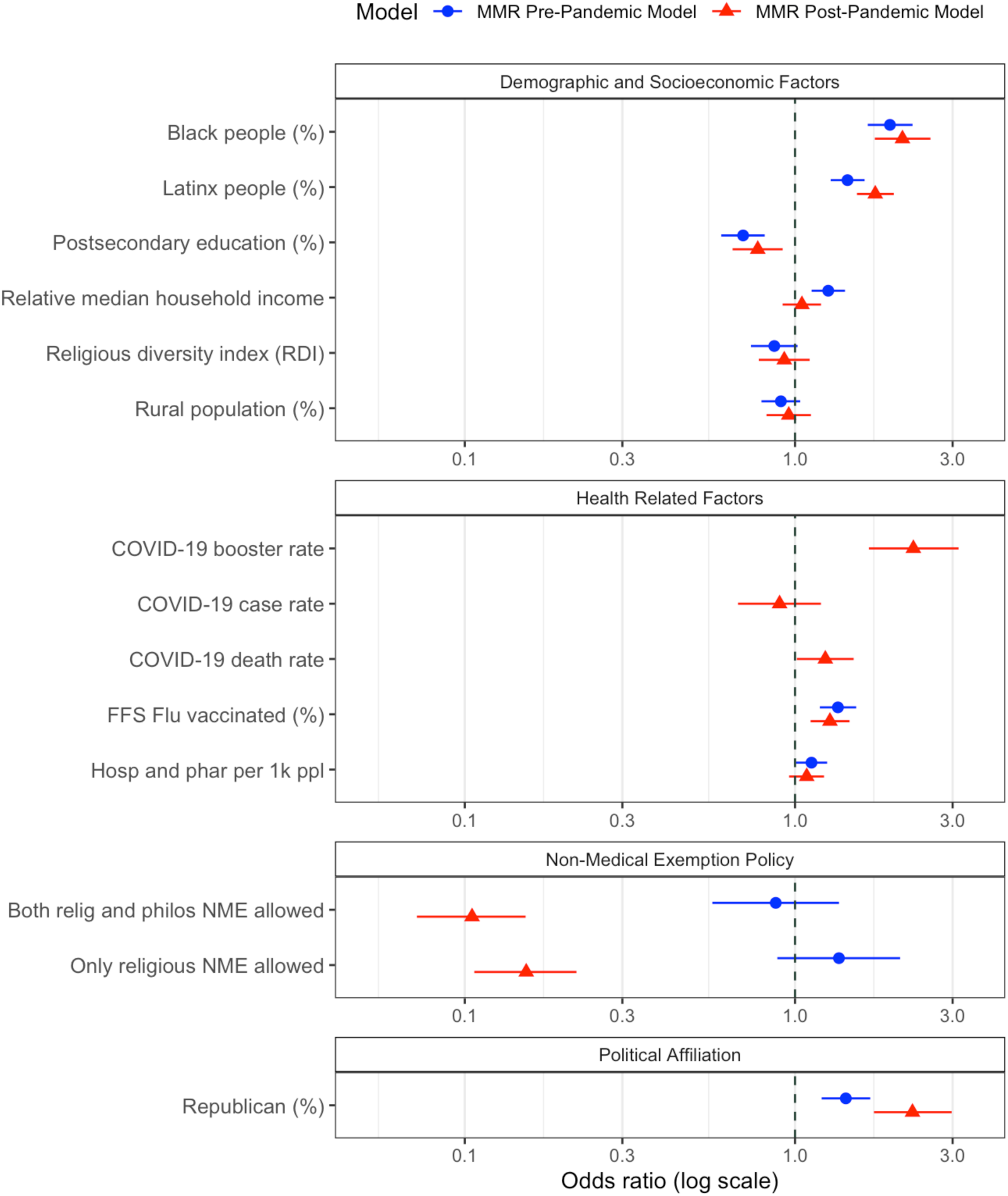
Results for the MMR Pre-Pandemic Model and MMR Post-Pandemic Model. The relationship between each explanatory factor and the likelihood of a county to have low MMR vaccination rates is presented as odds ratios. The red triangles and blue circles represent the odds ratios, while the corresponding lines indicate the 95% confidence intervals. An OR > 1 suggests that higher values of the factor are associated with counties maintaining MMR vaccination rates above 95%, whereas an OR < 1 suggests higher values are associated with counties having rates below 95%.

The MMR Post-Pandemic Model yields similar results to the MMR Pre-Pandemic Model, with a few key differences. After the COVID-19 pandemic, the associations with percentages of Black and Latinx populations, postsecondary education rates, FFS flu vaccinated percentage, and percentage of voters supporting Donald Trump in the 2020 presidential election remain consistent with those prior to the pandemic. However, certain factors significant in the pre-pandemic model, such as relative median household income and healthcare accessibility, become insignificant. Critically, state-level NME policies, which are insignificant in the pre-pandemic model, become the most significantly associated factor with low MMR rates post-pandemic, with the dual religious and philosophical NMEs slightly more significant than religious NMEs alone. Additionally, COVID-19 booster and death rates are positively associated with county-level MMR vaccination rates above 95% post-pandemic. However, our sensitivity analysis indicates that the association between COVID-19 death rates and MMR vaccination rates is more sensitive to threshold variation and should therefore be interpreted with caution. The complete results of the sensitivity analysis used to evaluate the 95% threshold are presented in Supplementary Section C.

### Factors associated with declining MMR vaccination rates during the pandemic

Results from the MMR Declining Model are presented in Figure 4 and Table S5. Consistent with the results of the MMR Post-Pandemic Model, the MMR Declining Model indicates that counties permitting non-medical exemptions (religious only or both philosophical and religious) are more likely to have declines in MMR vaccination rates during the pandemic. Counties with a higher rural population percentage and a high Religious Diversity Index (RDI) were also associated with declines. Conversely, counties with a higher relative median household income and a greater percentage of Latinx population were less likely to experience declines in MMR rates. We note that both the MMR Pre-Pandemic Model and the MMR Declining Model suggest that counties with a higher relative median household income are less likely to have low or declining rates. However, this association is insignificant in the MMR Post-Pandemic Model. Results for the sensitivity analysis for this model are presented in Supplementary Section C.

**Figure 4.**
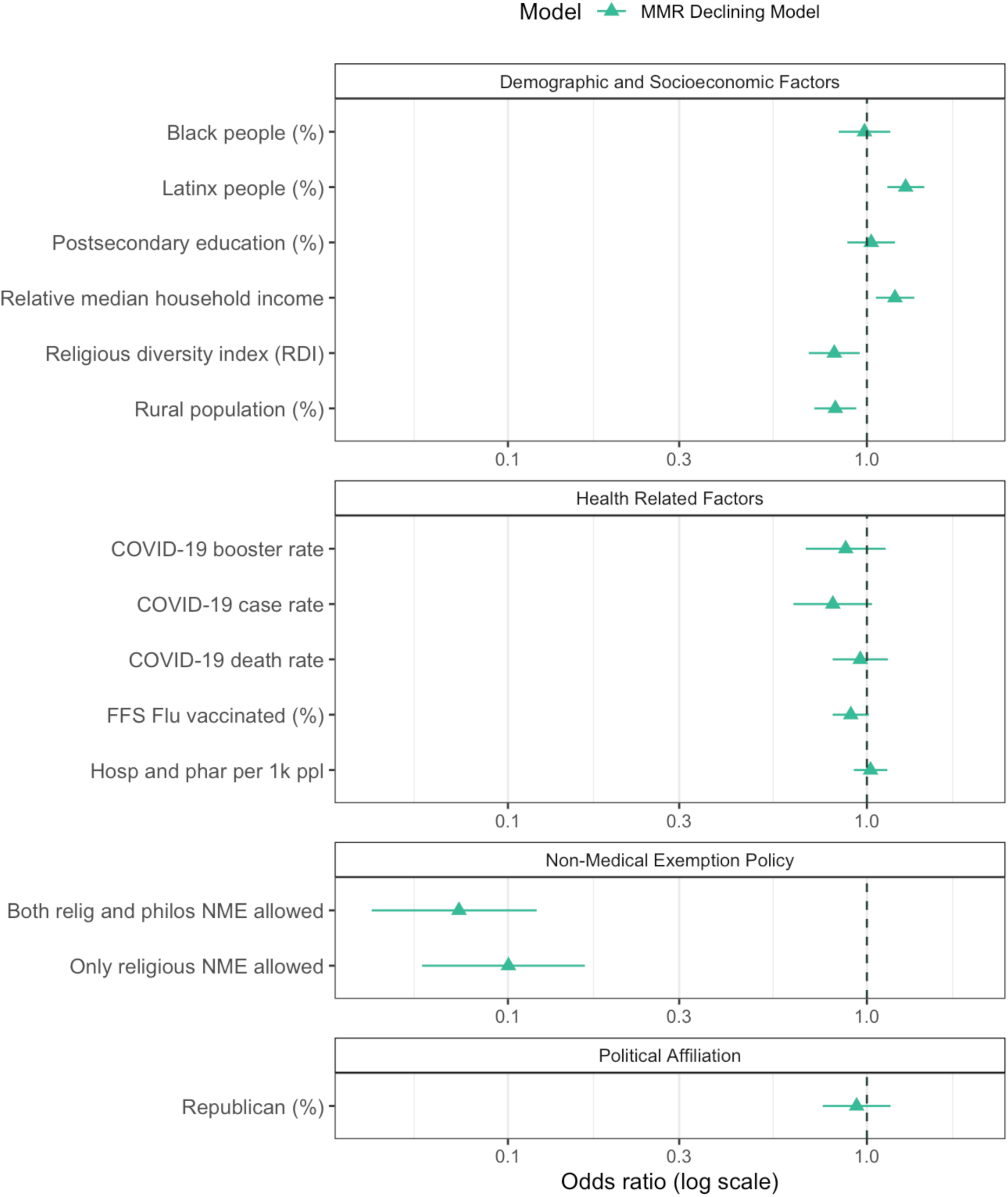
Results for the MMR Declining Model are presented as odds ratios, illustrating the relationship between each explanatory factor and the likelihood of a county to have substantially declining MMR vaccination rates, where a substantial decline is defined as those counties with MMR rate declines equal to or greater than the national county-level average of 2.36%. Green triangles represent the odds ratio, while the corresponding lines indicate the 95% confidence intervals. An OR > 1 suggests that counties with higher values of the factor are less likely to experience declining vaccination rates. Conversely, an OR < 1 suggests that counties with higher values of the factor are more likely to experience declining vaccination rates.

## Discussion

### Observed nationwide decline in MMR vaccination rates after the COVID-19 pandemic

Our county-level dataset complements the state and national-level CDC data,^4^ confirming a widespread decline in MMR vaccination rates in the U.S. following the COVID-19 pandemic, while critically revealing the significant heterogeneity in vaccination patterns both within and across states. By creating an open, centralized, high-resolution dataset, we offer a valuable resource to further explore the vaccination landscape in the U.S., and its implications for vaccine preventable disease.

### Demographic disparities reflected in MMR vaccination patterns

We found that counties with higher percentages of Black or Latinx populations were less likely to have low MMR vaccination rates both before and after the pandemic. Our findings may be attributed to mandatory childhood vaccination requirements in most U.S. schools, which has likely influenced MMR vaccination across diverse demographic groups,^26^ and helped maintain high vaccination rates in counties with large Black and Latinx populations, despite a historical tendency for these groups to exhibit stronger opposition to vaccinations and other healthcare interventions.^27,28^ While research from pre-pandemic periods lacks consensus on these associations,^7^ likely due to the spatial clustering of low MMR rates in various distinct communities, our findings align with a 2011 CDC report indicating that, since 2007, disparities in MMR vaccination coverage between non-Hispanic white children and racial or ethnic minorities have been virtually nonexistent.^29^

Furthermore, a higher percentage of Latinx population was less likely to be associated with MMR vaccination rate declines, while the Black population percentage showed no significant association, reinforcing the CDC’s reporting and suggesting these populations did not alter their MMR vaccination seeking behavior during the pandemic.

### Socioeconomic heterogeneity reflected in MMR vaccination patterns

Our analysis found that counties with a higher percentage of individuals with postsecondary education were more likely to have low MMR vaccination rates both before and after the pandemic, aligning with previous literature suggesting that highly educated parents may be more skeptical about vaccines or have stronger personal beliefs influencing their vaccination decisions for their children.^30^

Additionally, we observe that counties with a higher rural population percentage were more likely to experience significant declines during the pandemic. Several factors may contribute to this association. First, the current vaccine distribution system may not have adequately ensured sufficient vaccine availability in rural areas, especially amid the disruption of healthcare services caused by COVID-19. Second, differences in how rural residents perceived the politicized pandemic may have led to a stronger distrust in vaccine safety compared with more urban regions.^31^

Our results also indicate that higher Religious Diversity Index (RDI) values are associated with larger declines in county-level MMR vaccination rates, suggesting religiously diverse communities were increasingly likely to seek vaccine exemptions during the pandemic. These findings complement existing literature that points to the complex relationship between vaccine coverage and religious beliefs, due to drastically divergent attitudes towards vaccinations across religious groups.^32^ Additionally, past measles outbreaks in the U.S. have been linked to insular religious minority groups, particularly those with historically low vaccination uptake. We recommend further study of this association in the post-pandemic context.

Counties with higher relative median household incomes were less likely to have low MMR rates before the pandemic and less likely to have significant declines in MMR rates during the pandemic. This aligns with past literature linking lower-income households to lower vaccination rates,^33^ though, this association is not universally observed.^7^ Additionally, counties with higher healthcare accessibility – measured by number of hospitals and pharmacies per 1000 people - were less likely to have low MMR rates pre-pandemic, suggesting that limited access to public health facilities may impede childhood vaccination. However, this association was not significant in the MMR Post-Pandemic or MMR Declining Model, suggesting that low or declining MMR rates post-pandemic are better explained by other variables.

### Evidence for vaccine acceptance spillover

Our analysis reveals positive associations between acceptance of different types of vaccines, namely Flu, COVID-19, and MMR vaccines, contributing to the growing literature on general vaccine confidence patterns. The positive association between MMR vaccination rates and COVID-19 booster rates is unsurprising, suggesting that individuals who proactively seek non-mandatory vaccinations, i.e., COVID-19 booster, have greater overall trust in vaccine efficacy and safety, and are thus more likely to complete the recommended childhood vaccination series for their children. The additional association of MMR rates with flu vaccine rates reflects the more general spillover effects in vaccine acceptance across different routine vaccination types. These findings align with existing literature,^34^ and imply that increasing public trust in one vaccine type might increase trust in vaccinations more broadly.

### Contrasting role of political affiliation on MMR and COVID-19 vaccination behavior

Our results highlight a divergence in the relationship between population-level political affiliation and MMR vaccination rates compared to COVID-19 vaccination rates in the U.S. Previous research revealed a significant association between the percentage of Republican voters in a county with anti-vaccine attitudes and lower COVID-19 vaccination uptake.^12,35^ However, in this study, we observe that counties with a higher proportion of Republican voters in the 2020 presidential election were less likely to have low MMR vaccination rates both before and after the pandemic. Furthermore, the sensitivity analysis presented in Supplementary Section C confirms that this relationship remains consistent when using 2016 and 2024 election data for the Pre- and Post-Pandemic Models, respectively. One potential explanation of these results is because MMR is part of a long-established immunization program reinforced by mandatory school requirements,^8,36^ whereas the COVID-19 vaccine was a newly introduced vaccine (for all age groups), utilized novel technology, and was closely tied to a highly politicized pandemic – likely contributing to the disproportionate vaccine hesitancy.^12,35^ While these population-level findings contrast with post-pandemic survey data from Zhou et al, which suggests that Republican parents exhibit greater childhood vaccine hesitancy compared with Democrats and that county-level political affiliation does not show significant association with MMR vaccine hesitancy,^37^ in general, the relationship between political affiliation and childhood vaccination hesitancy remains inconclusive. Some studies link higher hesitancy to Republicans,^38^ while others point to data indicating greater hesitancy for Democrats,^39^ and some find no meaningful association at all.^7^ These inconsistent findings are likely due to the complexity of vaccination hesitancy, differences across vaccine-related decisions as well as distinctly different study designs. Given the inconsistencies in literature and the results of our study, we argue that, unlike COVID-19 vaccine hesitancy, political affiliation may not be a primary driver of childhood vaccine hesitancy directly. We conclude that addressing MMR vaccine hesitancy requires a bipartisan approach, focusing on strategies and policies that transcend political divisions to ensure equitable efforts in maintaining and improving childhood vaccination rates across various communities.

### Non-medical exemption policies reduce MMR vaccination rates

Our findings point to a notable shift in the role of NME policies during and after the pandemic, which becomes the predominant association with low MMR rates in the Post-Pandemic and Declining Models, reinforcing concerns about declining vaccine coverage under current vaccine exemption policies^20,40^. The phenomena are even more impactful in states with both religious and philosophical exemptions than those with only religious exemption. As shown in Figure S2 and Table S4, MMR vaccination rates declined on average 2.78% (from 94.46% to 91.68%) among counties in states allowing religious exemptions and 2.64% (93.02% to 90.38%) in those allowing both philosophical and religious exemptions, both greater than the national average decline. In stark contrast, counties in states with no NME policies experienced a modest increase of 0.33% (from 94.81% to 95.14%). Although the subset of counties differs slightly between analyses, the overall trend remains consistent. Furthermore, three of the four states that removed religious, philosophical, or both types of non-medical exemption options during the study period - Connecticut, Maine, and New York - experienced an increase in median county-level MMR rates. These findings further underscore the role of NME policies on MMR rates post-pandemic, and suggest that increasing the prevalence of NME options in the U.S. will further reduce vaccination coverage, and therefore increase the risk of measles outbreaks.^8,23,36,41^ These findings combined with past literature underscore the urgent need for evidence-based policy reform to protect public health while balancing religious and individual beliefs.^40,41^

### Limitations

While this study provides valuable insights into the current vaccination landscape and factors influencing MMR vaccination patterns in the U.S., it is subject to limitations. Firstly, the data is incomplete, as some states do not report county-level MMR vaccination data, limiting our nationwide analysis to 37 states. Second, the lack of standardized reporting for childhood vaccinations results in inconsistencies in reporting across states and thus affects the quality of the data available. For example, certain regions report their 2-dose MMR rates as part of a broader childhood vaccination series. Consequently, the MMR rate we assign for these locations represents a lower bound on the true MMR coverage. Third, the 2-dose MMR vaccination rate, which reflects the rate of kindergartners with complete measles vaccine records, may be low not only due to high vaccine refusal rates but also because of missing student records or delays in receiving the booster due to pandemic-related disruptions, among other factors. These factors introduce bias into the dataset; for example, counties with high immigrant populations may have a higher number of students with missing records, resulting in MMR rates that are lower than the actual rates. Fourth, the county-level percentage of flu-vaccinated FFS Medicare enrollees (65+) represent a subgroup of the population, and not likely to be the same group making the parental vaccination choices modeled in this study. Fifth, we represent the active state-level NME policies from 2017 (pre-pandemic) and 2022 (post-pandemic), which do not account for the delayed effect any policy changes may have on local vaccine behavior. Additionally, this analysis is purely statistical and does not reveal causal associations between the set of factors considered and MMR rates. Finally, this study is designed to improve our understanding of MMR vaccination uptake behavior; while MMR vaccination rates are directly related to the risk of measles, mumps and rubella outbreaks, we do not directly evaluate the risks or harms posed by these diseases.

## Conclusion

We provide a novel, high-resolution and comprehensive data set on MMR vaccination coverage in the U.S, spanning the entirety of the COVID-19 pandemic. The study revealed a significant nationwide decline in MMR vaccination rates following the pandemic and uncovers critical associations between MMR coverage and critical driving factors, in particular the role of non-medical exemption (NME) policies in reducing vaccination coverage during and after the COVID-19 pandemic. Our findings highlight the need for evidence-based vaccine policy, targeted vaccination interventions, and implementing culturally sensitive outreach programs to increase vaccine coverage in the U.S.

## Data Availability

All data collected for this analysis and their sources are available on GitHub at

https://github.com/CSSEGISandData/MMR_COVID_modeling

## Supplementary materials

### Data sharing statement

All data collected for this analysis and their sources are available on GitHub: https://github.com/CSSEGISandData/MMR_COVID_modeling.

### Section A: Data Collection and Processing

#### County-Level MMR Vaccination Rates

The focus of this study is to evaluate MMR vaccination coverage before and after the COVID-19 pandemic, thus we collected historical annual, county-level MMR vaccination data ranging from 2017-2024 for all U.S. counties providing this data. This section provides a detailed overview of the comprehensive MMR data collection process.

Where available, we collected county-level 2-dose MMR vaccination rates for kindergarten children by school year from each state’s Department of Health website for the study period. In some states this variable is not available, so the best available proxy was collected. The breakdown of this reporting is detailed in Supplementary Table S1.

In most cases, States released county-level vaccination coverage by school year for a range of vaccine types. When data was reported by calendar year, we assigned it to the closest school year based on state regulations. Kansas, New York and Michigan reported school-level MMR rates and Rhode Island reported city-level MMR rates (instead of county). For Kansas, New York and Michigan, we collected both the student enrollment statistics and the total vaccinated count for each school, aggregated both to county-level, and then computed county-level vaccination rates by dividing the county-level vaccinated count by the county-level kindergarten enrollment. For New York state, where not all school enrollments were available, we used the average MMR vaccination rate of all schools in a county as a proxy for the county-level rate. For Rhode Island vaccination rates at the city level were mapped to counties, and weighted by city population in 2020. West Greenwich and West Kingston are not included due to missing data. New Shoreham (2023-24) data is missing and adopts 2022-23 data. Warren (2020-21) data is missing and adopts 2021-22 data.

To evaluate the change in MMR vaccination rates before and after the pandemic, we define the pre-pandemic MMR vaccination rate as the average of the rates for the 2017-18, 2018-19, and 2019-20 school years, while the post-pandemic MMR rate is defined as the average of the rates for the 2022-23 and 2023-24 school years. If any county-level data is missing for a particular school year in this time period, data for that particular school year is omitted from the calculation of pre- or post-pandemic average MMR rates.

Our final dataset included county-level 2-dose MMR vaccination rates with both pre- and post-pandemic data for 37 states. These states include Alabama, Arizona, California, Colorado, Connecticut, Florida, Hawaii, Iowa, Kansas, Kentucky, Louisiana, Maine, Maryland, Massachusetts, Michigan, Minnesota, Mississippi, Missouri, New Jersey, New Mexico, New York, North Carolina, North Dakota, Oklahoma, Oregon, Pennsylvania, Rhode Island, South Carolina, South Dakota, Tennessee, Texas, Utah, Vermont, Virginia, Washington, Wisconsin, and Wyoming. In total, the MMR vaccination rates were collected in these 37 states.

Several states are excluded from our analysis due to MMR data limitations. Specifically, Washington D.C. is excluded because pre-pandemic MMR data is unavailable, and Montana is excluded because post-pandemic MMR data is unavailable. Alaska, Arkansas, Delaware, Idaho, Nebraska, Nevada, New Hampshire, Ohio, and West Virginia are excluded because only state-level data is publicly available. Georgia, Indiana and Illinois are excluded because they only report 1-dose MMR vaccination rates for children aged 19-35 months, which does not align with our focus on 2-dose MMR rates modeled in this study. These exclusions are necessary to reduce potential biases in the data and subsequent analysis. However, we chose to include data from South Carolina, Utah, Wisconsin, and Wyoming, despite their reporting of 2-dose MMR data for age groups slightly different from kindergarten. We also chose to include data for the 14 states that included the 2-dose MMR vaccine as part of a complete vaccine series (i.e. DTaP, Polio, VAR, HepA, HepB, PCV, etc.). While this data represents a lower bound on 2-dose MMR rates for these locations, we believe the inclusion of these states adds valuable information to our study without compromising the overall integrity of the analysis. Supplementary Table S1 provides detailed information on the collected MMR coverage data for each state.

#### Explanatory Factors

We collected publicly available data on demographics and socioeconomics, political affiliation, vaccine policy (NMEs), COVID-19 metrics and other health-related variables to uncover significant associations with MMR vaccination behavior. The datasets, including their sources and other key details, are described below. If data on any variable is missing for a county that county is excluded from the analysis. Counties with missing values in the explanatory factors were dropped from the model. In total, we have 2,266 counties in the study. We summarize all the explanatory factors and their sources in Table S2.

***Demographic and socioeconomic factors*** are collected at the county-level and include the percentage of Black and Latinx populations, the percentage of rural population; the percentage of people with postsecondary degrees or higher; relative household median income (ratio of county-level household median income to the state-level average); and religious diversity index (RDI). These variables were obtained from multiple sources including the U.S. Census Bureau,^1^ American Community Survey (ACS),^2^ CDC and Agency for Toxic Substances and Disease Registry (ATSDR),^3^ and Public Religion Research Institute (PRRI).^4^ Demographic and socioeconomic factors were selected based on their relevance to childhood vaccinations or their links to vaccine hesitancy or low vaccination rates in past research.^5^ We used the 2020 demographic data as it is the most recent census data available and the most relevant for our study period.

***Political affiliation*** was measured by the percentage of voters supporting the Republican candidate in the 2020 presidential election for each U.S. county. This variable is included because political beliefs have been shown to influence attitudes towards vaccinations.^6,7^ To ensure the robustness of our findings, we conducted a sensitivity analysis using data from the 2016 and 2024 presidential elections, presented in Figure S9 and S10. The data is available from the MIT Election Data and Science Lab.^8^

***Health-related factors*** that may influence MMR vaccination rates include COVID-19 metrics (health outcomes and vaccinations), healthcare accessibility, and vaccine uptake behavior for other diseases. We included various COVID-19 metrics, including COVID-19 cumulative case and death rates per 100,000 people as of June 30^th^, 2022,^9^ and cumulative COVID-19 booster rate as of March 3, 2022.^10,11^ We included COVID-19 case and death rates as a proxy for the severity of the pandemic in each county leading up to the study period (the 2022-2023 and 2023-2024 school years). We included booster rates to understand the relationship between optional COVID-19 vaccination behavior and MMR coverage, amid concerns that attitudes towards the COVID-19 vaccine may have influenced childhood vaccination rates.^12^

We used the number of hospitals and pharmacies per 1000 people as a proxy for healthcare accessibility.^13^ We included this variable as low healthcare accessibility was historically tied to lower MMR rates, particularly before measles was declared eliminated in the U.S. in 2000.^5^

Additionally, we used the county-level percentage of fee-for-service Medicare enrollees receiving annual flu vaccinations (FFS flu vaccinated percentage) as a proxy for flu-vaccination levels.^14^ Flu vaccination data was included to further explore county-level differences in uptake across various vaccine types. The pre-pandemic FFS flu vaccinated percentage was averaged from 2017, 2018 and 2019 data, while the post-pandemic percentage was based on the most recent 2021 data. While FFS Medicare flu vaccination data reflects a different population (65+) than childhood vaccinations, we believe it serves as a reasonable proxy for overall flu vaccination rates and captures a critical aspect of a populations vaccine seeking behavior.

***Non-medical exemption (NME) policies*** at the state level, which have been identified as potential facilitators of vaccine hesitancy,^5^ were included as a categorical explanatory variable. States are classified based on whether they allowed only medical exemptions, only religious exemptions, or both religious and philosophical exemptions. Original data was obtained from the CDC^15^ and updated based on reports from the National Conference of State Legislators (NCSL).^16^ We used NMEs from 2017 for the pre-pandemic period (2017-2019) and NMEs from 2022 for the post-pandemic period (2022-2024). In the pre-pandemic period, 35 out of 37 states permitted religious exemptions, while 14 allowed personal exemptions. In the post-pandemic period, these numbers decreased to 32 states and 12 states, respectively. Key policy changes that occurred during our study period included New York removing its religious exemption option in 2019, Washington removing its personal exemption option in 2019, Maine removing both religious and personal exemption options in 2019, and Connecticut removing its religious exemption option in 2021.^16^ Although Mississippi introduced a religious exemption option in April 2023, this change was not included in our analysis as it occurred after the start of the post-pandemic study period and was therefore not reflective of the trends in our model.

**Table S2.** Summary statistics and corresponding data sources of the target and explanatory variables.

| Factors | N | Mean | SD | Median | Min | Max | Source |
| --- | --- | --- | --- | --- | --- | --- | --- |
| **MMR Vaccination Rates** |  |  |  |  |  |  |  |
| Pre-pandemic MMR vaccination rates | 2266 | 0.94 | 0.06 | 0.95 | 0.30 | 1.00 | CDC and local health departments |
| Post-pandemic MMR vaccination rates | 2266 | 0.92 | 0.07 | 0.93 | 0.32 | 1.00 | CDC and local health departments |
| **Health Related Factors** |  |  |  |  |  |  |  |
| COVID-19 case rate | 2266 | 25956.43 | 5397.28 | 25861.08 | 9046.75 | 68110.44 | JHU CSSE COVID-19 dashboard |
| COVID-19 death rate | 2266 | 386.95 | 163.97 | 381.83 | 0.00 | 1222.95 | JHU CSSE COVID-19 dashboard |
| COVID-19 booster rate | 2266 | 23.38 | 8.43 | 22.04 | 5.25 | 56.85 | COVID-19 vaccination tracking project |
| Pre-pandemic FFS Flu vaccinated (%) | 2266 | 42.87 | 9.80 | 44.30 | 6.30 | 67.00 | County Health Rankings & Roadmaps (CHR&R) |
| Post-pandemic FFS Flu vaccinated (%) | 2266 | 44.94 | 10.65 | 46.00 | 5.00 | 71.00 | County Health Rankings & Roadmaps (CHR&R) |
| Hospitals and pharmacies per 1000 people | 2266 | 0.29 | 0.19 | 0.25 | 0.00 | 2.17 | County Health Rankings & Roadmaps (CHR&R) |
| **Demographic and Socioeconomic Factors** |  |  |  |  |  |  |  |
| Black people (%) | 2266 | 8.88 | 14.14 | 2.48 | 0.00 | 87.46 | U.S. Census Bureau |
| Latinx people (%) | 2266 | 11.21 | 15.29 | 5.11 | 0.34 | 97.68 | U.S. Census Bureau |
| Higher education (%) | 2266 | 54.00 | 10.72 | 54.00 | 7.40 | 90.30 | U.S. Census Bureau |
| Relative median household income | 2266 | 1.00 | 0.22 | 0.96 | 0.49 | 2.63 | U.S. Census Bureau |
| Rural population (%) | 2266 | 57.79 | 32.00 | 59.00 | 0.00 | 100.00 | U.S. Census Bureau |
| Religious diversity index (RDI) | 2266 | 0.63 | 0.13 | 0.65 | 0.23 | 0.90 | Public Religion Research Institute (PRRI) |
| **Political Affiliation** |  |  |  |  |  |  |  |
| Republican (%) | 2266 | 63.75 | 16.62 | 66.34 | 8.73 | 96.18 | MIT Election Data and Science Lab |
| **Non-Medical Exemption Policy** |  |  |  |  |  |  |  |
| NME1 (Only religious NME allowed) | 1144 | / | / | / | / | / | National Conference of State Legislatures (NCSL) |
| NME2 (Both relig and philos NME allowed) | 921 | / | / | / | / | / | National Conference of State Legislatures (NCSL) |

#### Detailed Non-Medical Exemption Data Analysis

In this section, we present a detailed analysis of non-medical exemption policies across all 50 states and D.C., along with a comparison of these policies to MMR vaccination rate distributions. Table S3 summarizes all state-level NME policies in the United States, while Figure S1 visually depicts their geographic distribution. Figure S2 and Table S4 depict the relationship between NME policies and MMR rates. These tables and figures reinforce key findings from the multivariable logistic regression model, which indicate that NME policies are strongly associated with low and declining MMR rates post-pandemic, with dual religious and philosophical NMEs more closely linked to low and declining MMR rates than religious NMEs alone.

**Table S3.**
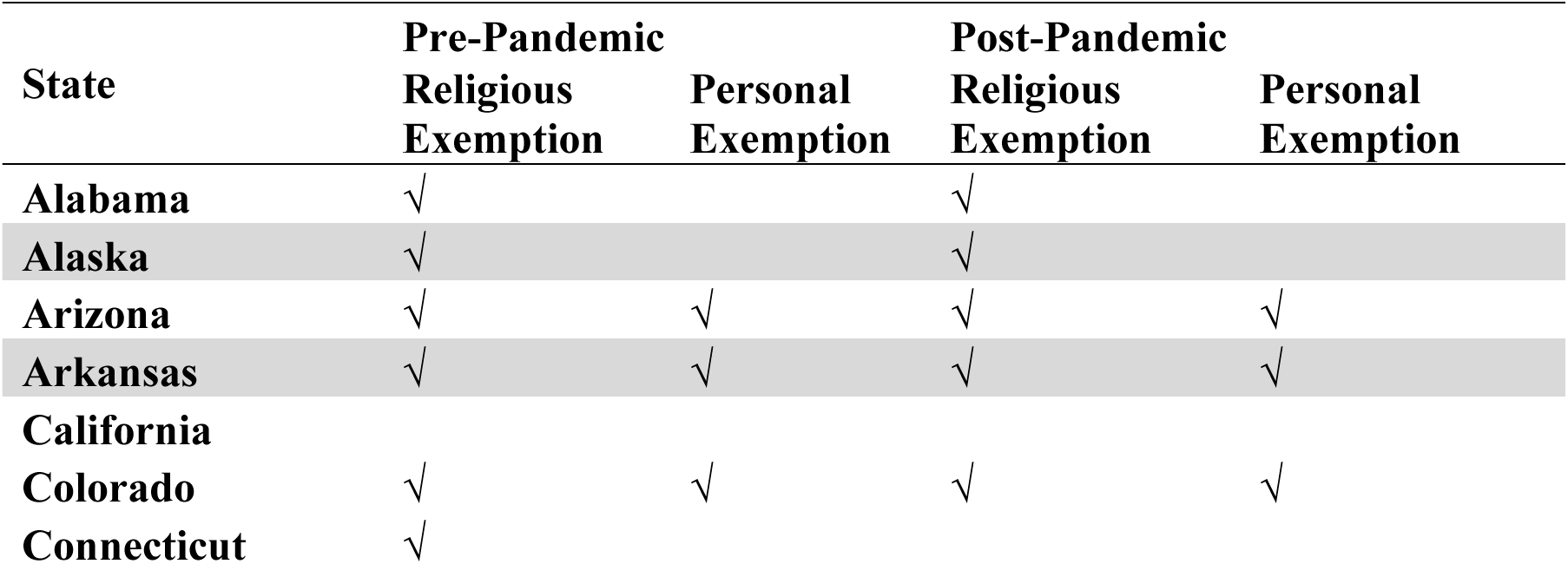
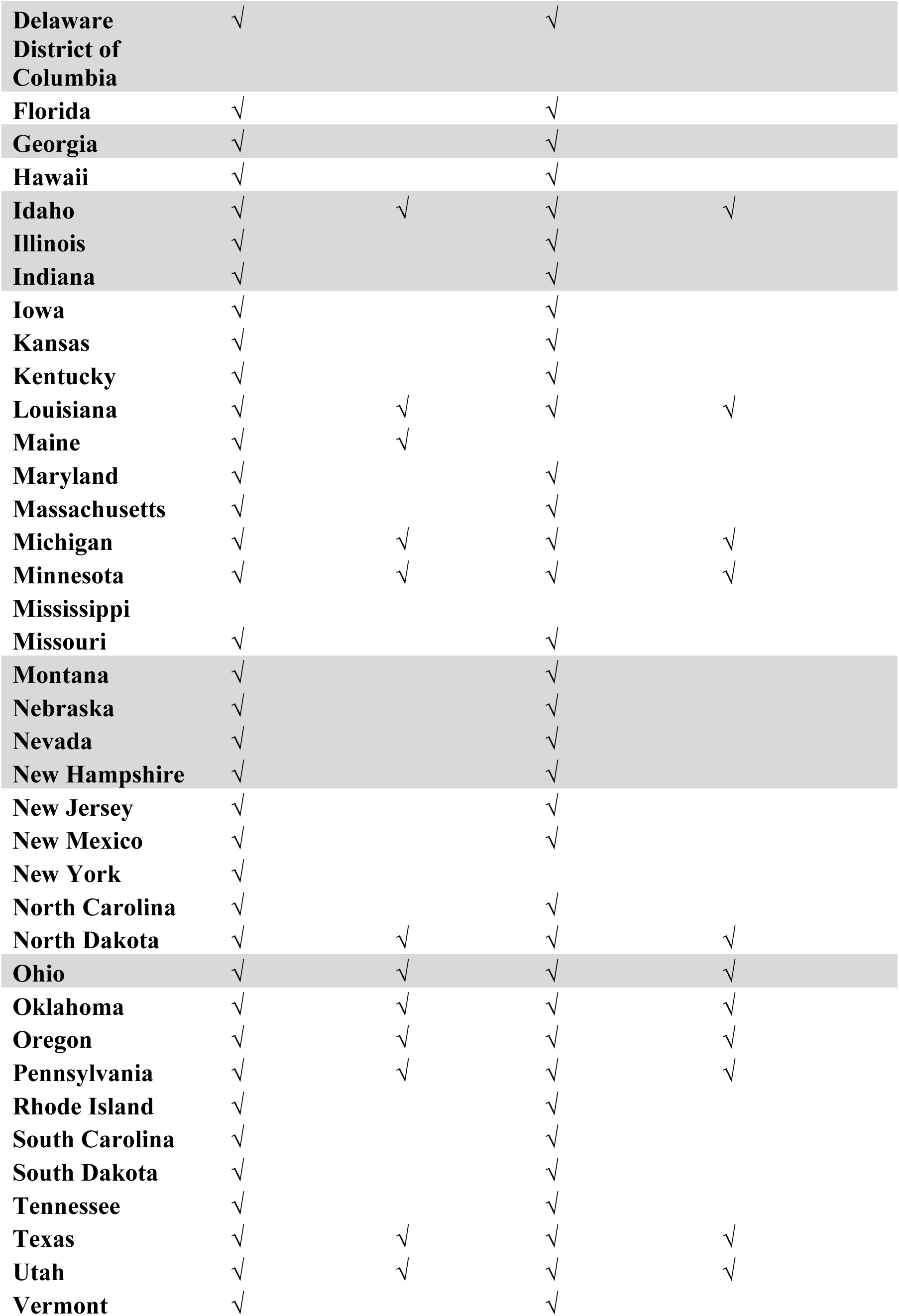
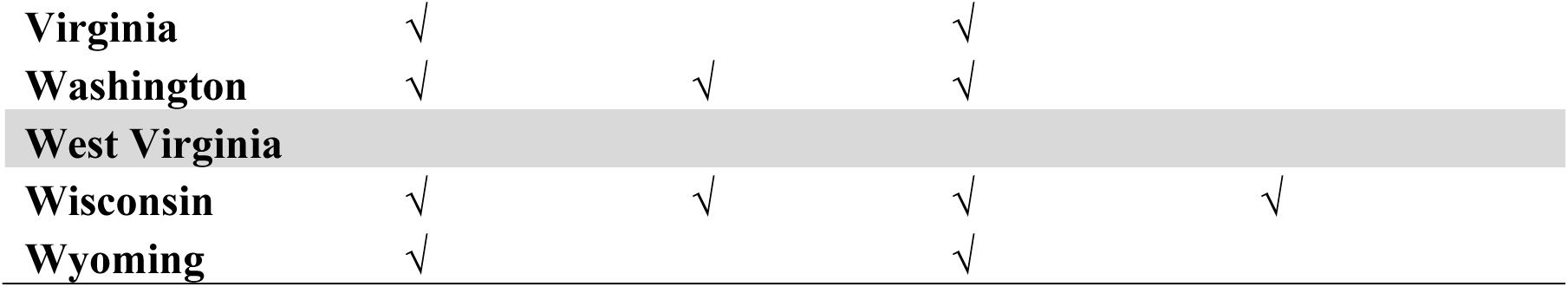
State-Level Non-Medical Exemption Policies in 2017 and 2022. The data was obtained from the CDC^15^ and updated using the NCSL data.^16^ The 2017 data represent pre-pandemic NME policies, while the 2022 data represent post-pandemic NME policies. All 51 state-level NME policies (including D.C.) are present in the table. States excluded from our study are highlighted grey.

**Figure S1:**
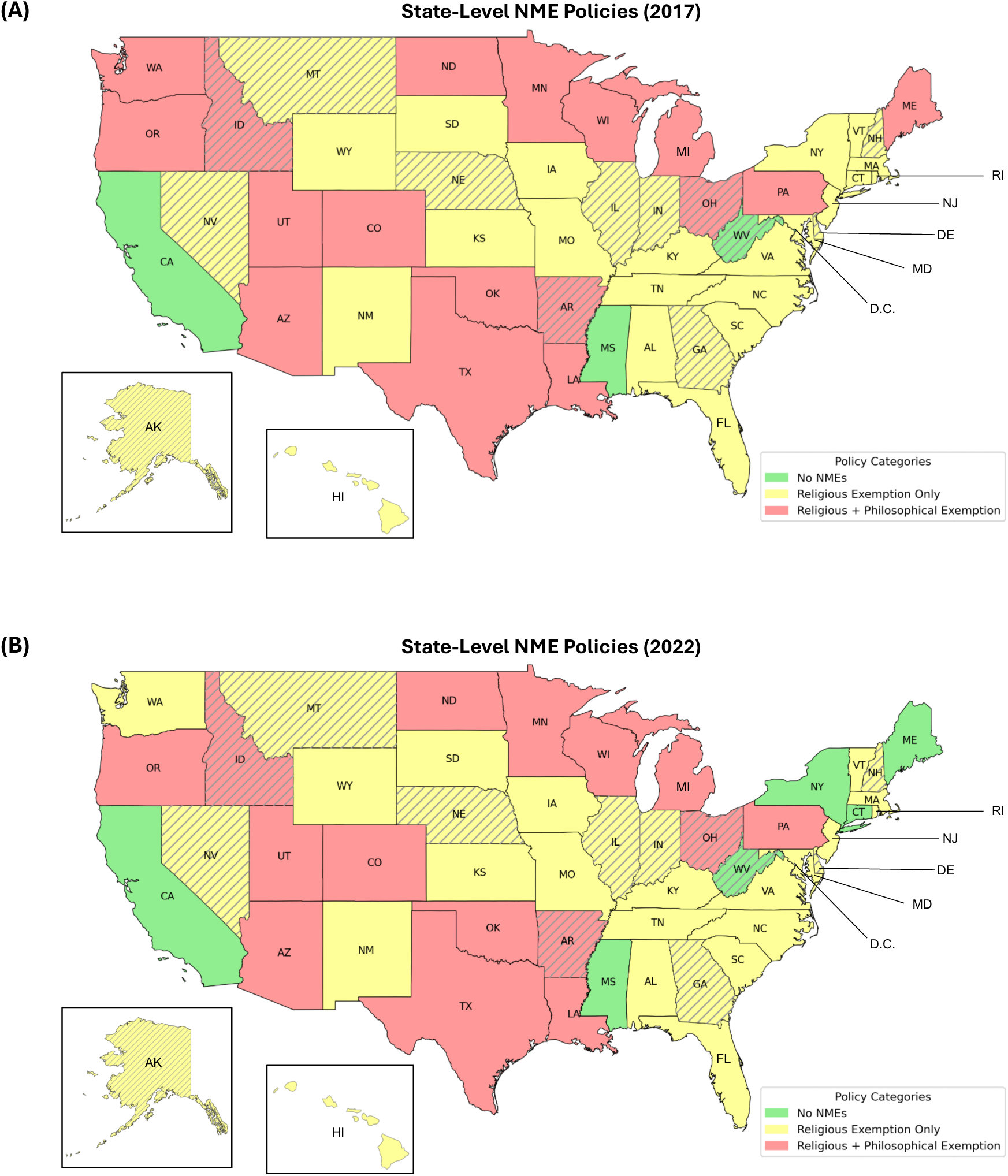
Maps representing state-level Non-Medical Exemption (NME) policies in the U.S. (A) depicts state-level NME policies in 2017, while (B) shows NME policies in 2022. These years were selected because they represent the start of our pre-pandemic and post-pandemic periods, respectively. States that are not included in our study are marked with grey diagonal lines (hatch patterns).

**Figure S2:**
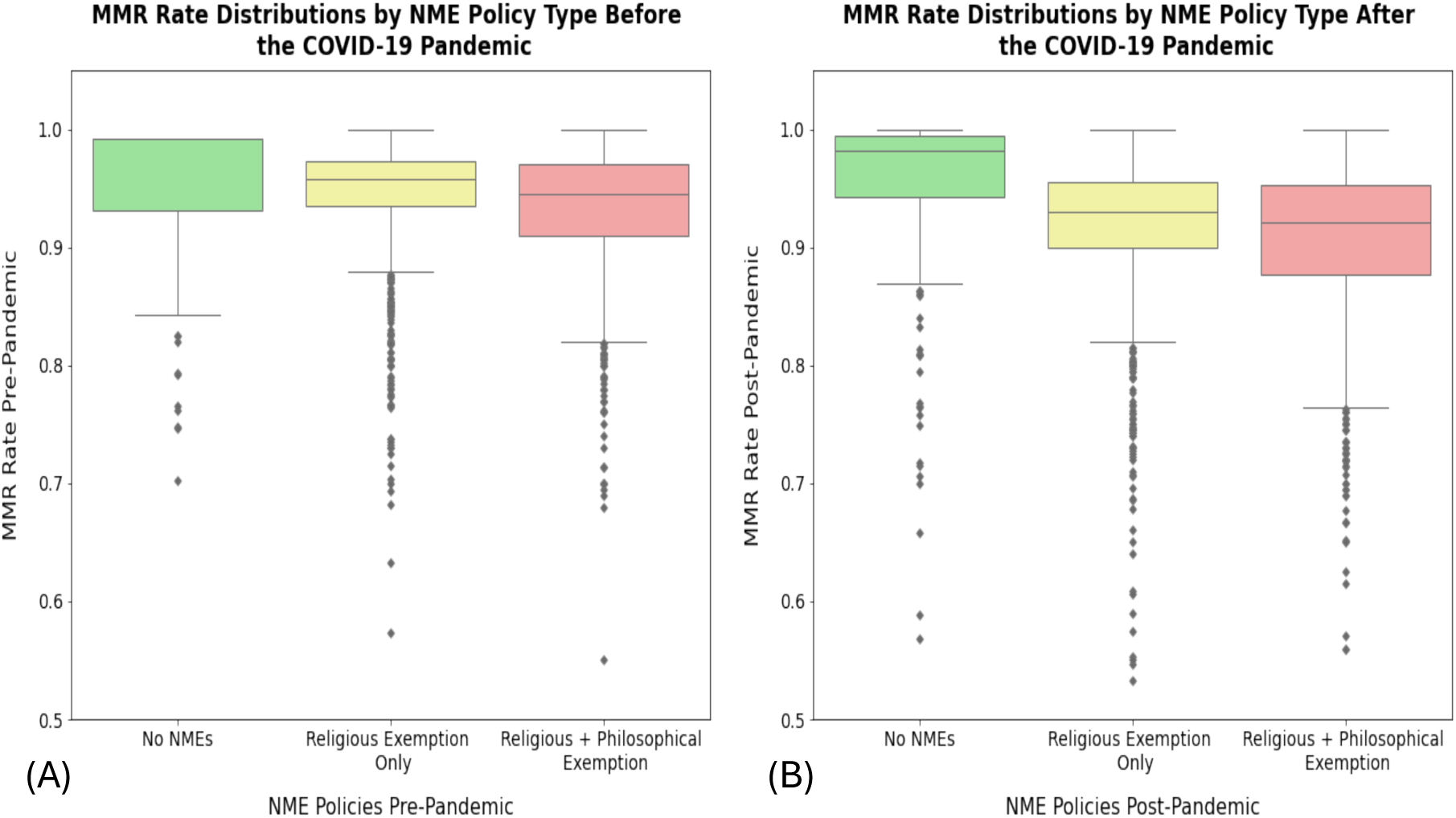
Distribution of MMR rates for counties included in our study, grouped by NME policy type. Panel (A) shows the pre-pandemic distribution, and panel (B) shows the post-pandemic distribution. A clear decline in MMR rates is observed in counties that allow religious exemptions or both religious and philosophical exemptions. This decline is notably more pronounced in the post-pandemic period compared to the pre-pandemic period.

**Table S4:** Documents MMR rate distributions for counties included in our study. This table accompanies Figure S2 and highlights the significant post-pandemic decline in MMR rates, particularly in counties permitting religious exemptions or both religious and philosophical exemptions.

| **State-Level**<br>**NME Policies** | **Number of**<br>**Counties**<br>**Pre-**<br>**Pandemic** | **Average**<br>**County-**<br>**Level**<br>**MMR**<br>**Rate Pre-**<br>**Pandemic** | **Median**<br>**County-**<br>**Level**<br>**MMR**<br>**Rate Pre-**<br>**Pandemic** | **Number of**<br>**Counties**<br>**Post-**<br>**Pandemic** | **Average**<br>**County-**<br>**Level**<br>**MMR**<br>**Rate Post-**<br>**Pandemic** | **Median**<br>**County-**<br>**Level**<br>**MMR**<br>**Rate**<br>**Post-**<br>**Pandemic** |
| --- | --- | --- | --- | --- | --- | --- |
| No NMEs | 139 | 94.81% | 99.20% | 225 | 95.14% | 98.17% |
| Religious<br>Exemptions<br>Only | 1206 | 94.46% | 95.74% | 1175 | 91.68% | 93.00% |
| Religious and<br>Philosophical<br>Exemptions | 921 | 93.02% | 94.54% | 866 | 90.38% | 92.07% |
| All Policies<br>(Total) | 2266 | 93.89% | 95.33% | 2266 | 91.53% | 93.08% |

### Section B: Detailed Methods Description

#### MMR Pre-Pandemic Model and MMR Post-Pandemic Model

We employed multivariable weighted logistic regression to identify factors associated with low MMR vaccination rates before and after the pandemic. We chose 95% as the threshold, as it is widely used in academic publications and government policies to refer to the minimum MMR vaccine target for herd immunity.^17^ To examine low MMR vaccination coverage, we dichotomized the target variables (pre- and post-pandemic MMR vaccination rates) into counties with MMR vaccination rates > 95% and those with rates ≤ 95%. We then fit two separate logistic regression models, one for pre-pandemic (MMR Pre-Pandemic Model) and one for post-pandemic (MMR Post-Pandemic Model) MMR vaccination rates, using counties with rates ≤ 95% as the reference baseline. We conducted an extensive sensitivity analysis to validate the robustness of our results for this threshold, evaluating MMR rates at 94.0%, 94.5%, 95.0%, 95.5%, and 96.0%. As presented in Figure S6 and Figure S7, our findings confirm that the results remain stable across these thresholds. We define the two separate logistic regression models for the pre-pandemic and post-pandemic periods below:

1) MMR Pre-Pandemic Model (counties with low MMR rates pre-pandemic as the binary target variable):

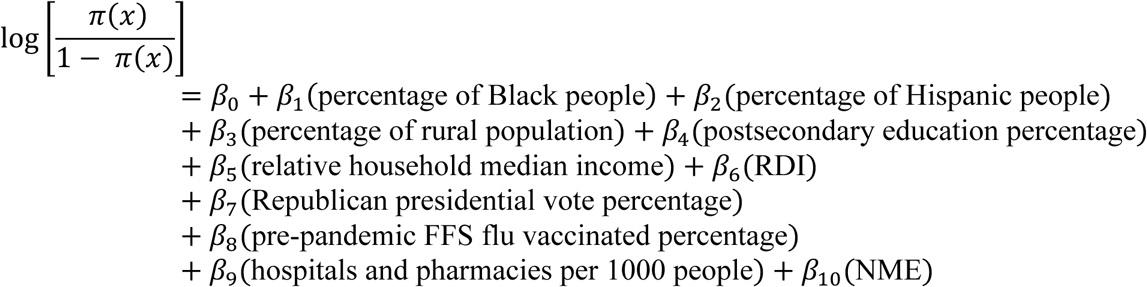

2) MMR Post-Pandemic Model (counties with low MMR rates post-pandemic as the binary target variable):

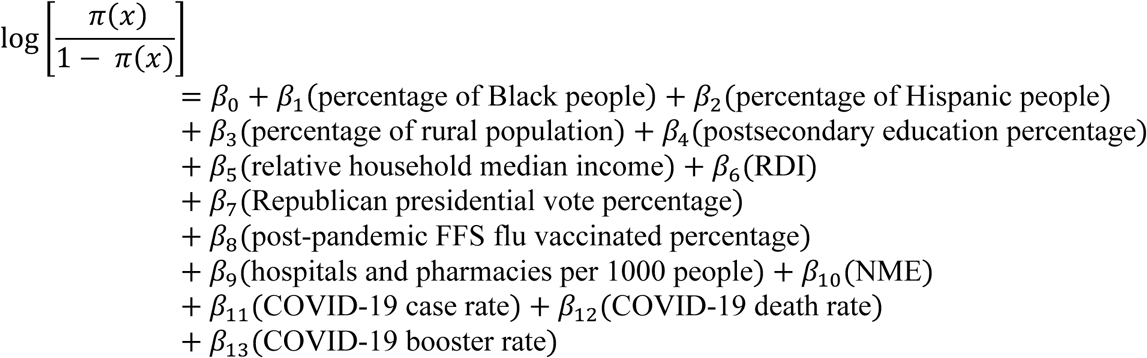

where π(*x*) represents the probability of a county having an MMR vaccination rate more than 95% before or after the pandemic, and 1 − π(*x*) represents the probability of a county having an MMR vaccination rate less than or equal to 95% before or after the pandemic. Counties with an MMR vaccination rate equal to or below the 95% threshold pre- or post-pandemic serve as the baseline reference group for the MMR Pre-Pandemic Model and MMR Post-Pandemic Model, respectively. Figure S3A and S3B present maps that visually depict the geographic distribution of counties in these baseline reference groups for both models. Both models are weighted by county population to account for differences in population size across counties: *w_i_* = log (*pop_i_*).

Each model presents the relationship between explanatory variables and the outcome as odds ratios, along with their corresponding 95% confidence intervals. The odds is represented by 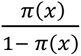. An odds ratio (OR) < 1 indicates counties are more likely to have vaccination rates less than or equal to 95%, an OR > 1 indicates counties are less likely to have vaccination rates below 95%, and OR = 1 suggests no association.

The MMR Pre-Pandemic Model does not include variables for COVID-19 health outcomes or COVID-19 booster rate. However, because the MMR Post-Pandemic Model aims to identify factors associated with low MMR vaccination rates after the pandemic, we included additional COVID-19 related factors into the model. All independent variables are continuous, except for the non-medical exemption (NME) policy, which has three categories: no non-medical exemption policies permitted (NME 0), counties permitting only religious exemptions (NME 1), and counties permitting both religious and philosophical exemptions (NME 2). This allows for an examination of the impact of different exemption policies on MMR vaccination rates. Furthermore, we note that the inclusion of NMEs makes this a multi-scale model, with all variables implemented at the county-level except for NMEs, which are applied at the state level.

#### MMR Joint Model

We also develop a combined logistic regression model that integrates pre- and post-pandemic MMR rates into a single analysis with interaction terms to further reinforce the findings of our MMR Pre-Pandemic and MMR Post-Pandemic Models.

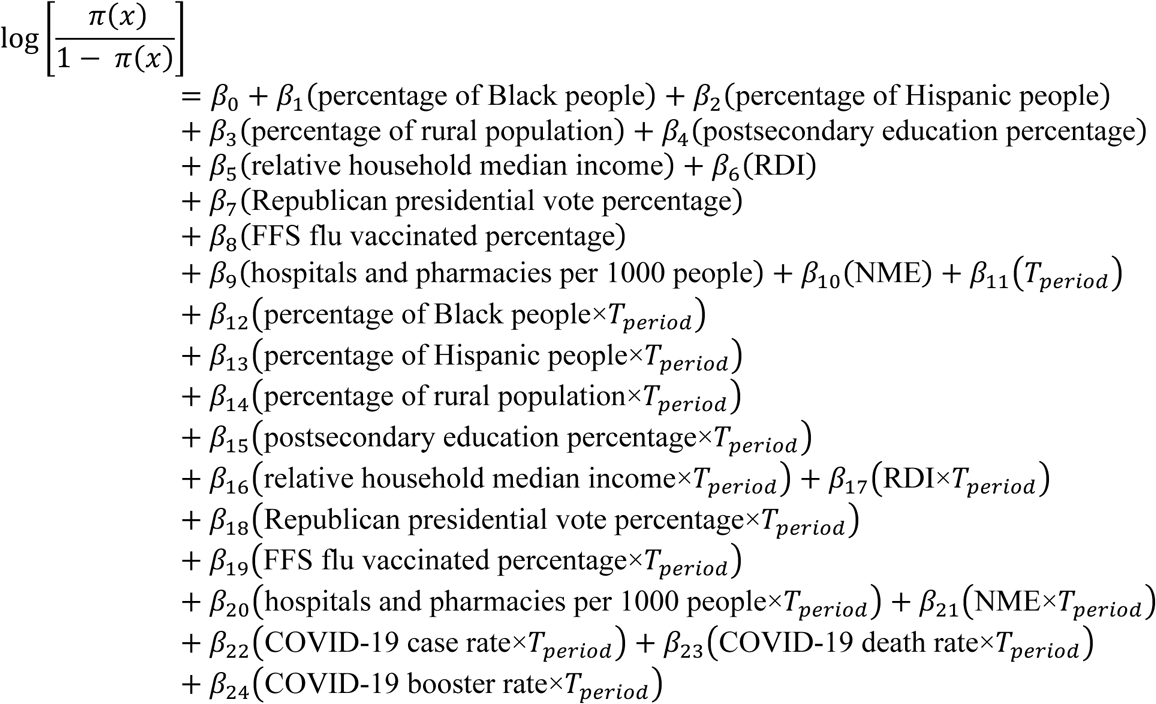

where π(*x*) represents the probability of a county having an MMR vaccination rate more than 95% before or after the pandemic, and 1 − π(*x*) represents the probability of a county having an MMR vaccination rate less than or equal to 95% before or after the pandemic. *T_period_* serves as a time variable, where *T_period_* = 0 during the pre-pandemic period and *T_period_* = 1 during the post-pandemic period. COVID-19 related factors are set as 0 when *T_period_* = 0. Counties with an MMR vaccination rate below 95% pre- or post-pandemic serve as the baseline reference group for the MMR Joint Model. This model is also weighted by county population to account for differences in population size across counties: *w_i_* = log (*pop_i_*).

#### MMR Declining Model

To investigate factors associated with the declining county-level MMR vaccination rates during the pandemic, we fit an additional model, the MMR Declining Model. This model used the same set of explanatory variables as the MMR Post-Pandemic Model, with the dependent variable adjusted to compare counties with substantial declines in MMR vaccination rates during the pandemic. A county was classified as a substantial decline or “declining” if its MMR vaccination rate decreased by -2.36% or more, aligning with the average county-level decline from the pre-pandemic to post-pandemic periods. In line with our Pre-Pandemic and Post-Pandemic models, we conducted a sensitivity analysis on this threshold, evaluating MMR rate declines of -1.5%, - 2.0%, -2.5%, -3.0%, -3.5%, and -4.0%. Our findings indicate that the results remain relatively stable, as detailed in Figure S8. The number of counties classified as “declining” based on the selected threshold of -2.36% is 977 (where the total number of counties with any reported decline is 1,688). The MMR Declining Model is described below:

MMR Declining Model (with declining counties as the binary target variable):

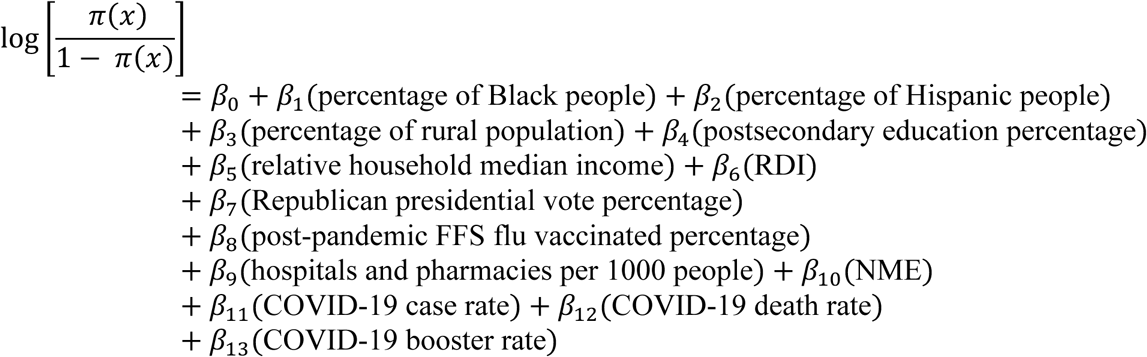

Counties with declining MMR vaccination rates during the pandemic represent the baseline reference group for the MMR Declining Model, where 1 − π(*x*) represents the probability that a county experienced declining MMR vaccination rates and π(*x*) represents the probability that a county did not experience declining MMR vaccination rates. Figure S3C presents a map that visually depicts the geographic distribution of counties in the baseline reference group for the MMR Declining Model. The model is weighted by county population to account for differences in population size across counties: *w_i_* = log (pop*_i_*).

The MMR Declining Model presents the relationship between explanatory variables and the outcome as odds ratios, along with their corresponding 95% confidence intervals. For statistically significant factors, an odds ratio greater than 1 (OR > 1) indicates that counties are less likely to have declining MMR vaccination rates. Conversely, an odds ratio less than 1 (OR < 1) suggests that counties are more likely to have declining MMR rates. An OR = 1 suggests no association. All variables in this model, except for the target variable, are identical to those in the MMR Post-Pandemic Model.

**Figure S3.**
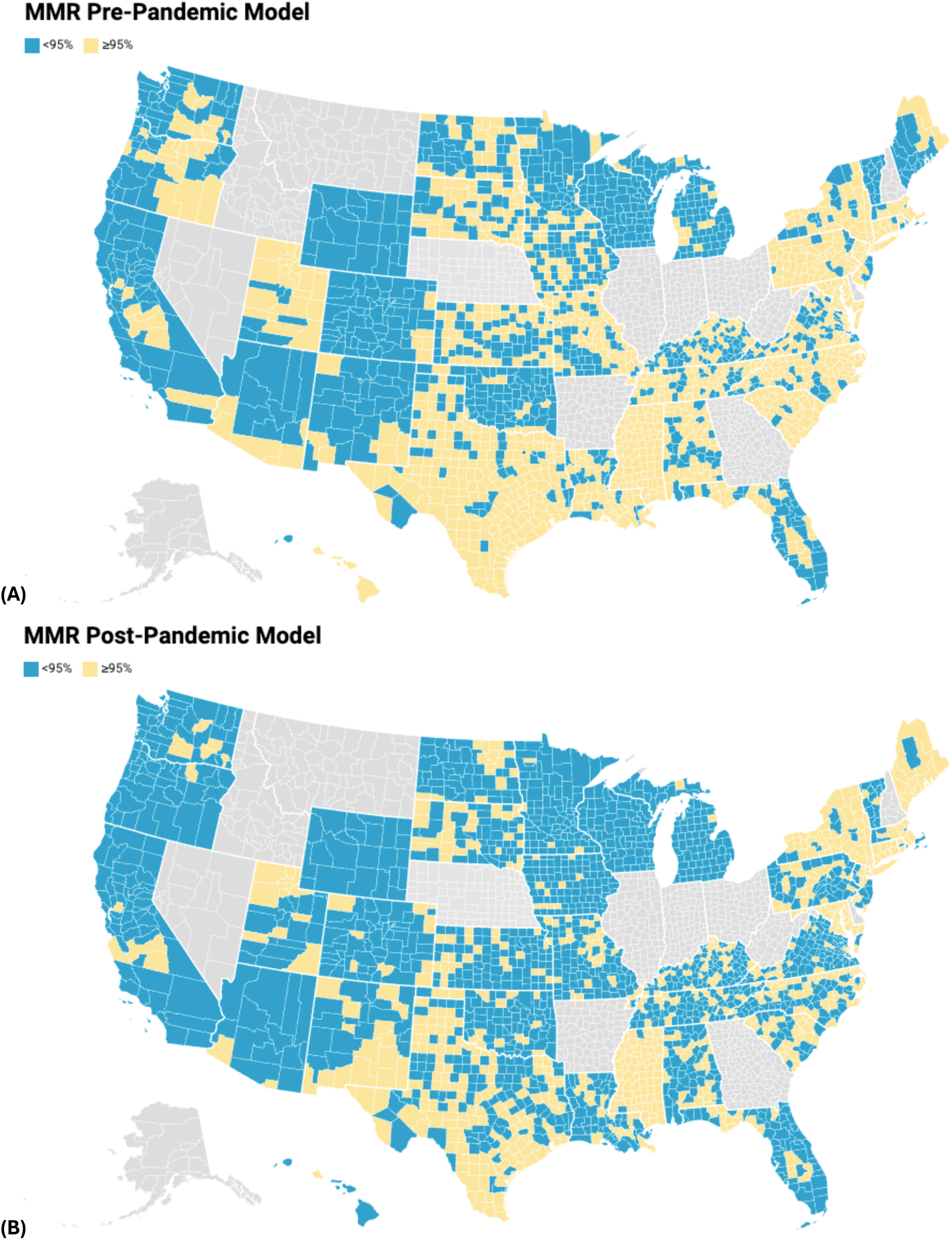
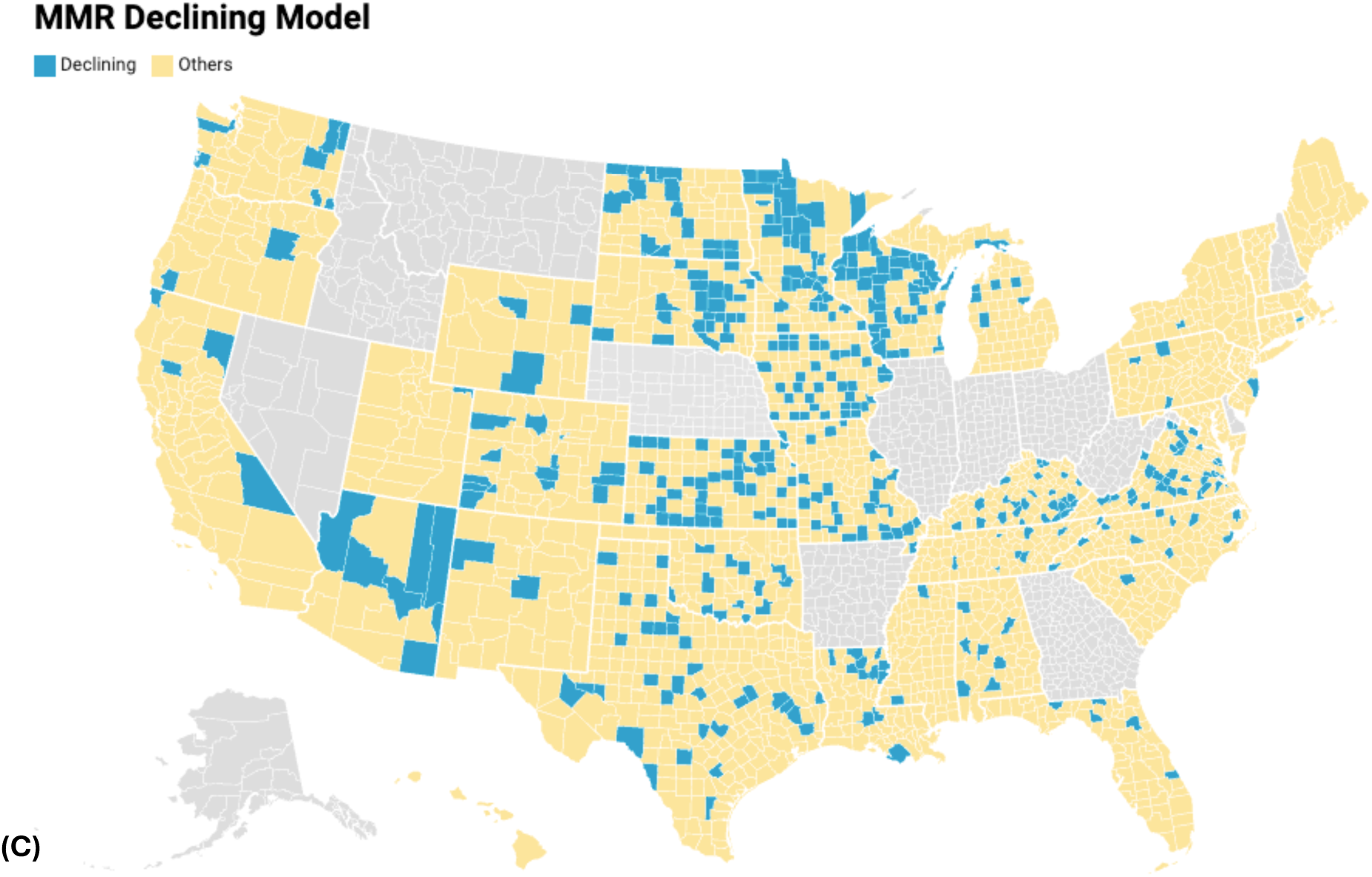
Maps illustrating the baseline reference groups (colored in blue) for the binary target variables of each multivariable weighted logistic regression model. Panel (A) displays the counties selected as the baseline reference group for MMR Pre-Pandemic Model (pre-pandemic MMR vaccination rates lower than or equal to 95%). Panel (B) shows the counties selected as the baseline reference group for MMR Post-Pandemic Model (post-pandemic MMR vaccination rates lower than or equal to 95%). Panel (C) presents the counties selected as the baseline reference group for MMR Declining Model (MMR vaccination rate declines greater than or equal to -2.36%, matching the average county-level decline). Counties in grey are those with missing MMR data. Maps are created by Datawrapper.^18^

#### Model Validation

In this section, we present visualizations of the data – including a correlation matrix and assessments of linearity assumptions, to support the use of a logistic regression framework for modeling this problem. We conclude that our modeling approach is well-suited to addressing this problem.

**Figure S4.**
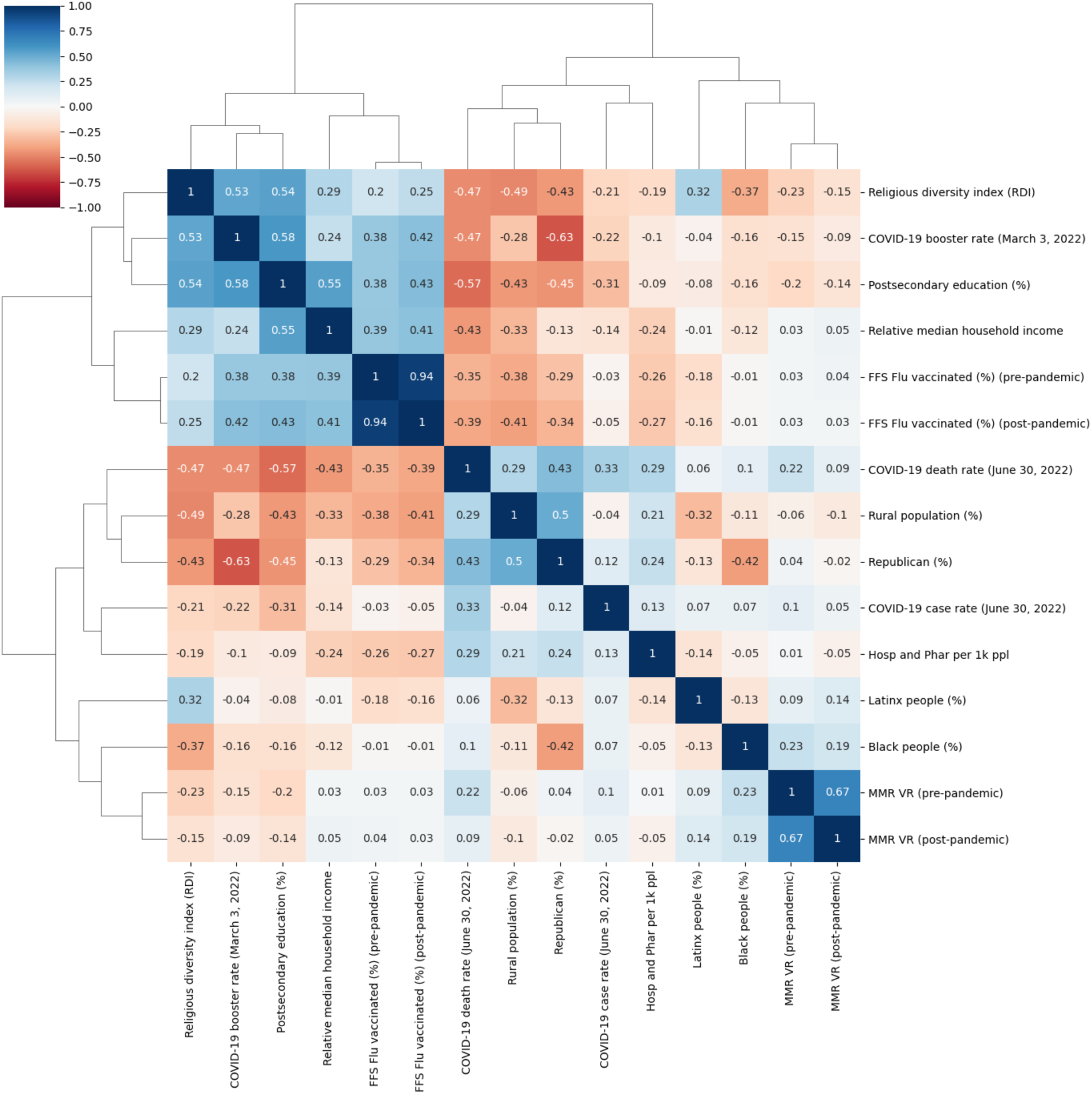
Correlation matrix of the studied variables. A Pearson correlation matrix of all variables used in the logistic regression models. Variables with an absolute correlation of 0.75 or higher were excluded to reduce multicollinearity.

**Figure S5.**
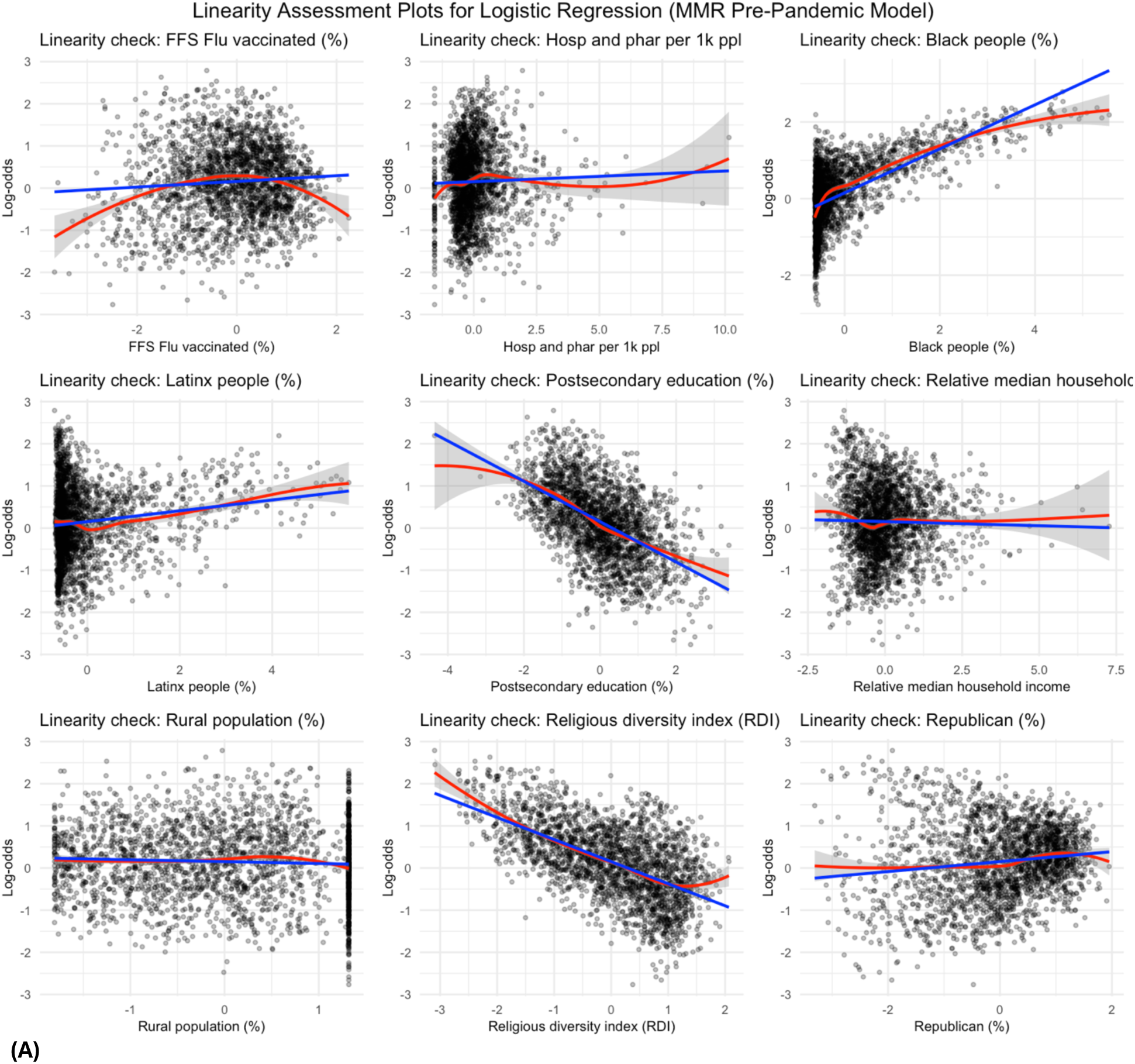
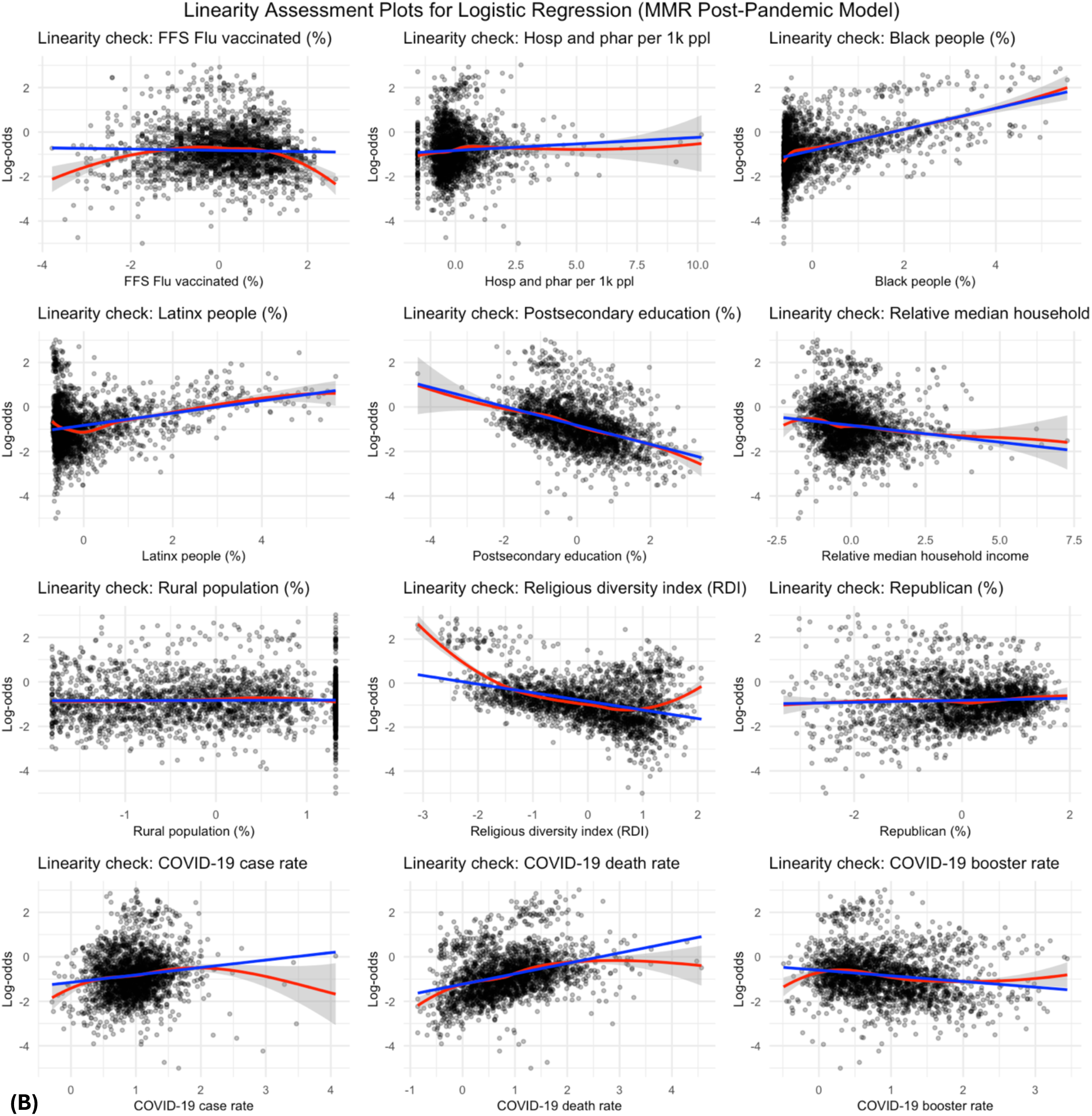
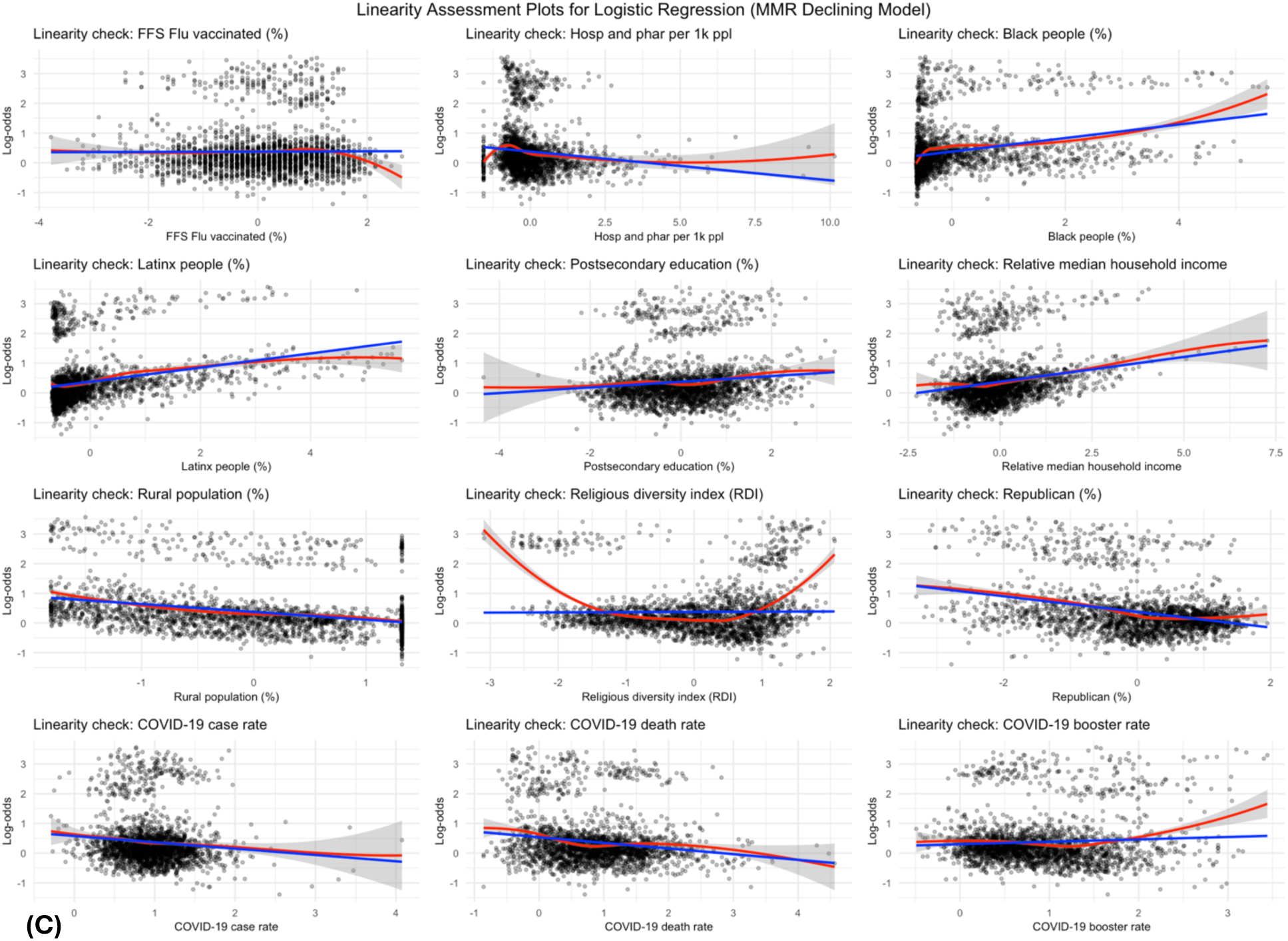
Charts illustrating the linearity assumption in logistic regression, where input data is assumed to be linearly related to the log-odds of the dependent variable. Panel (A) presents the linearity assumption for the Pre-Pandemic Model, Panel (B) presents the linearity assumption for the Post-Pandemic Model, and Panel (C) presents the linearity assumption for the Declining Model.

### Section C: Detailed Model Results and Sensitivity Analysis

This section provides additional detail on the model results by presenting the complete numerical outputs for each model. To further demonstrate the robustness of the findings from the multivariable logistic regression model, we conducted a comprehensive sensitivity analysis for each selected threshold. Additionally, we performed a sensitivity analysis on the elections data, evaluating our findings using the respective elections results from 2016 and 2024. Finally, we present the results of our joint logistic regression model to further reinforce the findings from our independent logistic regression models.

#### Model Results

Table S5 presents the odds ratios (ORs), 95% confidence intervals (CI), and statistical significance for all explanatory variables, complementing the results presented in Figure 3 (MMR Pre-Pandemic and MMR Post-Pandemic Models) and Figure 4 (MMR Declining Model). All results align with the corresponding figures.

For the MMR Pre-Pandemic and MMR Post-Pandemic Models, the relationship between each explanatory factor and the likelihood of a county to have low MMR vaccination rates is presented as an odds ratio. An OR > 1 suggests that higher values of the factor are associated with counties maintaining MMR vaccination rates > 95%, whereas an OR < 1 suggests higher values are associated with counties having rates ≤ 95%. For the MMR Declining Model, the odds ratios reflect the relationship between each explanatory factor and the likelihood that a county experienced a substantial decline in MMR vaccination rates, defined as a decline greater than the national county-level average of 2.36%. An OR > 1 suggests that counties with higher values of the factor are less likely to experience declining vaccination rates. Conversely, an OR < 1 suggests that counties with higher values of the factor are more likely to experience declining vaccination rates.

**Table S5:** Results for logistic regression models. Table S5 details the outputs of the 3 logistic regression models, the MMR Pre-Pandemic, MMR Post-Pandemic, and MMR Declining Models. This table presents the odds ratios (ORs), 95% confidence intervals (CI), and statistical significance for all explanatory variables. This table serves as a companion for Figure 3 and Figure 4.

| Models | MMR Pre-Pandemic Model |  |  | MMR Post-Pandemic Model |  |  | MMR Declining Model |  |  |
| --- | --- | --- | --- | --- | --- | --- | --- | --- | --- |
| Factors | OR | 95% CI | p-values | OR | 95% CI | p-values | OR | 95% CI | p-values |
| **Health Related Factors** |  |  |  |  |  |  |  |  |  |
| COVID-19 case rate |  |  |  | 0.90 | (0.67, 1.20) | 0.465 | 0.80 | (0.62, 1.03) | 0.090 |
| COVID-19 death rate |  |  |  | 1.24* | (1.01, 1.51) | 0.036 | 0.96 | (0.80, 1.14) | 0.637 |
| COVID-19 booster rate |  |  |  | 2.28*** | (1.67, 3.13) | <0.001 | 0.87 | (0.68, 1.13) | 0.299 |
| Flu vaccinated (%) | 1.35*** | (1.19, 1.53) | <0.001 | 1.28*** | (1.11, 1.46) | <0.001 | 0.90 | (0.80, 1.01) | 0.082 |
| Hospital and pharmacy count per 1000 people | 1.12* | (1.01, 1.25) | 0.038 | 1.08 | (0.96, 1.22) | 0.189 | 1.02 | (0.92, 1.14) | 0.670 |
| **Demographic and Socioeconomic Factors** |  |  |  |  |  |  |  |  |  |
| Black people (%) | 1.94*** | (1.66, 2.27) | <0.001 | 2.11*** | (1.74, 2.57) | <0.001 | 0.98 | (0.83, 1.16) | 0.854 |
| Latinx people (%) | 1.44*** | (1.28, 1.62) | <0.001 | 1.75*** | (1.54, 1.99) | <0.001 | 1.28*** | (1.14, 1.45) | <0.001 |
| Postsecondary education (%) | 0.70*** | (0.60, 0.81) | <0.001 | 0.77** | (0.65, 0.92) | 0.004 | 1.03 | (0.88, 1.20) | 0.723 |
| Relative median household income | 1.26*** | (1.12, 1.42) | <0.001 | 1.05 | (0.92, 1.20) | 0.482 | 1.20** | (1.06, 1.36) | 0.004 |
| Rural population (%) | 0.91 | (0.79, 1.04) | 0.153 | 0.96 | (0.82, 1.12) | 0.580 | 0.82** | (0.71, 0.93) | 0.003 |
| Religious diversity index (RDI) | 0.87 | (0.74, 1.02) | 0.081 | 0.93 | (0.78, 1.11) | 0.407 | 0.81* | (0.69, 0.96) | 0.012 |
| **Political Affiliation** |  |  |  |  |  |  |  |  |  |
| Republican (%) | 1.42*** | (1.20, 1.69) | <0.001 | 2.27*** | (1.73, 2.98) | <0.001 | 0.94 | (0.75, 1.16) | 0.552 |
| **Non-Medical Exemption Policy** |  |  |  |  |  |  |  |  |  |
| NME1 (Only religious NME allowed) | 1.36 | (0.88, 2.08) | 0.160 | 0.15*** | (0.11, 0.22) | <0.001 | 0.10*** | (0.06, 0.16) | <0.001 |
| NME2 (Religious and personal NME allowed) | 0.87 | (0.56, 1.36) | 0.550 | 0.11*** | (0.07, 0.15) | <0.001 | 0.07*** | (0.04, 0.12) | <0.001 |

Significance label: <0.001: \*\*\*; <0.01: \*\*; <0.05: \*

#### Sensitivity Analysis of MMR Pre-Pandemic Threshold

Our MMR Pre-Pandemic Model examines factors associated with low county-level MMR rates, defined as rates equal to or below the commonly used 95% threshold, which is widely accepted as the minimum target for achieving herd immunity. ^17^ In Figures S6 and Table S6, we present sensitivity analyses for this threshold for the Pre-Pandemic Model, to evaluate the robustness of our findings. Overall, our findings confirm the consistency and stability of the model outputs.

**Figure S6.**
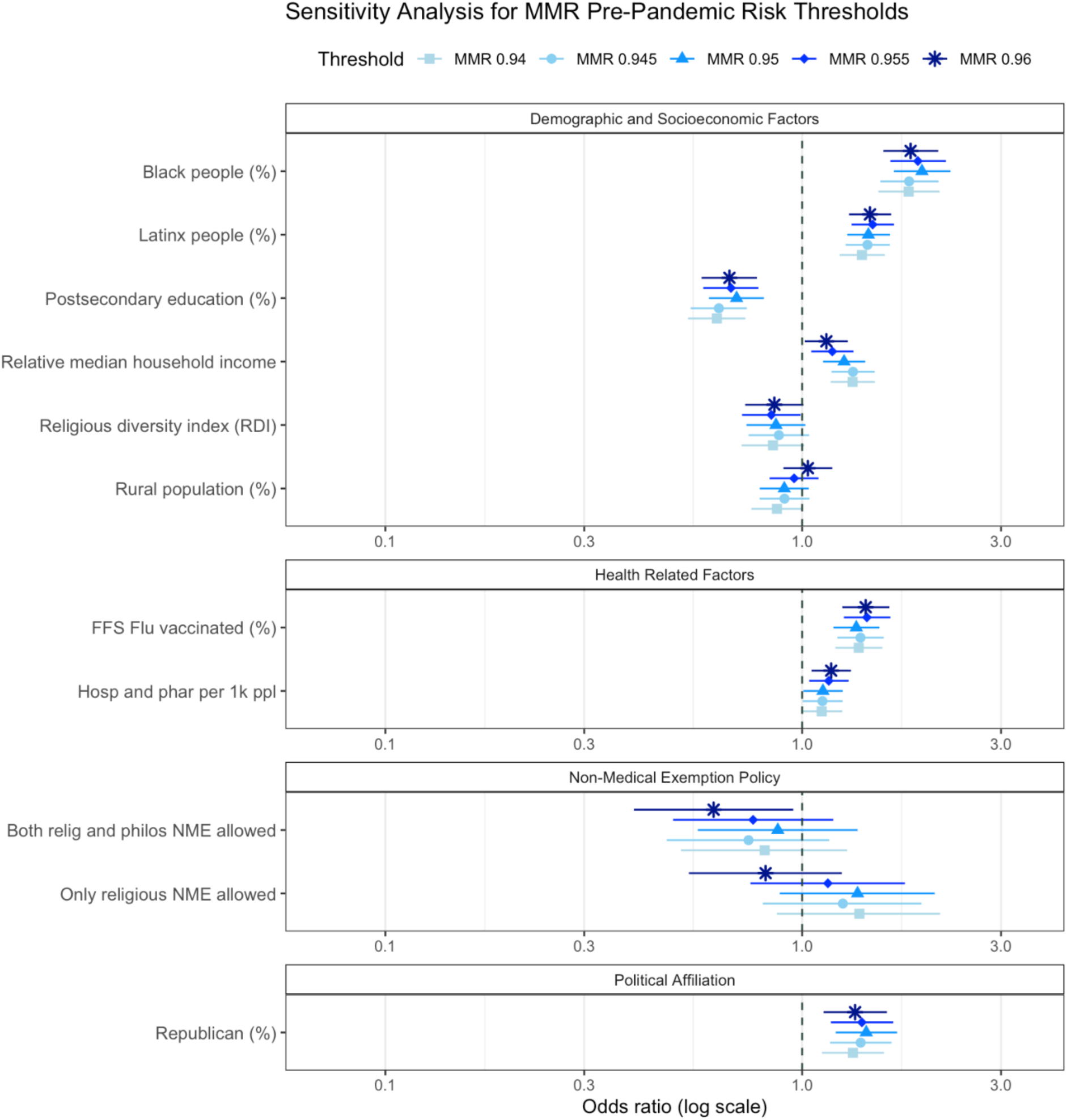
Results of the sensitivity analysis for the MMR Pre-Pandemic Model. This figure illustrates the relationship between each explanatory factor and MMR vaccination rate as odds ratios, evaluating the following MMR thresholds: 94.0%, 94.5%, 95.0%, 95.5%, and 96.0%. The odds ratio of each threshold is represented by a corresponding shape in the legend, while the colored lines indicate the 95% confidence interval. An OR > 1 suggests that higher values of the factor are associated with counties maintaining MMR vaccination rates above the given threshold, whereas an OR < 1 suggests higher values are associated with counties having rates equal to or below the given threshold. The size of the baseline reference group for each threshold (i.e., the number of counties below each threshold) is as follows: 826 (94.0%), 917 (94.5%), 1057 (95%), 1177 (95.5%), 1308 (96.0%). Our findings confirm the consistency and robustness of the results.

**Table S6:** Presents the sensitivity analysis for the MMR Pre-Pandemic Model, reporting odds ratios, 95% confidence intervals, and p-values for each explanatory factor. This table serves as a companion to Figure S6 and documents the relationship between each factor and MMR vaccination rates, evaluating the following MMR thresholds: 94.0%, 94.5%, 95.0%, 95.5%, and 96.0%. An OR > 1 suggests that higher values of the factor are associated with counties maintaining MMR vaccination rates above the given threshold, whereas an OR < 1 suggests higher values are associated with counties having rates equal to or below the given threshold. Statistical significance is denoted as follows: p <0.05 (*), p < 0.01 (**), p < 0.001 (***). The size of the baseline reference group for each threshold (i.e., the number of counties below each threshold) is as follows: 826 (94.0%), 917 (94.5%), 1057 (95%), 1177 (95.5%), 1308 (96.0%). Our findings confirm the consistency and robustness of the results.

| Characteristic | Pre Model (0.94) |  |  | Model (0.945) |  |  | Model (0.95) |  |  | Model (0.955) |  |  | Model (0.96) |  |  |
| --- | --- | --- | --- | --- | --- | --- | --- | --- | --- | --- | --- | --- | --- | --- | --- |
|  | OR<sup>†</sup> | 95% CI | p-value | OR<sup>†</sup> | 95% CI | p-value | OR<sup>†</sup> | 95% CI | p-value | OR<sup>†</sup> | 95% CI | p-value | OR<sup>†</sup> | 95% CI | p-value |
| FFS Flu vaccinated (%) | 1.37*** | 1.20, 1.56 | <0.001 | 1.38*** | 1.22, 1.57 | <0.001 | 1.35*** | 1.19, 1.53 | <0.001 | 1.43*** | 1.26, 1.63 | <0.001 | 1.42*** | 1.25, 1.62 | <0.001 |
| Hosp and phar per 1k ppl | 1.11 | 1.00, 1.25 | 0.058 | 1.12* | 1.00, 1.25 | 0.046 | 1.12* | 1.01, 1.25 | 0.038 | 1.16** | 1.04, 1.29 | 0.008 | 1.17** | 1.05, 1.31 | 0.004 |
| Black people (%) | 1.80*** | 1.52, 2.14 | <0.001 | 1.81*** | 1.54, 2.12 | <0.001 | 1.94*** | 1.66, 2.27 | <0.001 | 1.90*** | 1.63, 2.21 | <0.001 | 1.82*** | 1.57, 2.12 | <0.001 |
| Latinx people (%) | 1.39*** | 1.23, 1.58 | <0.001 | 1.43*** | 1.27, 1.62 | <0.001 | 1.44*** | 1.28, 1.62 | <0.001 | 1.48*** | 1.31, 1.66 | <0.001 | 1.45*** | 1.30, 1.64 | <0.001 |
| Postsecondary education (%) | 0.62*** | 0.53, 0.73 | <0.001 | 0.63*** | 0.54, 0.74 | <0.001 | 0.70*** | 0.60, 0.81 | <0.001 | 0.68*** | 0.58, 0.79 | <0.001 | 0.67*** | 0.57, 0.78 | <0.001 |
| Relative median household income | 1.32*** | 1.17, 1.50 | <0.001 | 1.32*** | 1.18, 1.49 | <0.001 | 1.26*** | 1.12, 1.42 | <0.001 | 1.18** | 1.05, 1.33 | 0.005 | 1.14* | 1.01, 1.29 | 0.027 |
| Rural population (%) | 0.87 | 0.76, 1.00 | 0.053 | 0.91 | 0.79, 1.04 | 0.165 | 0.91 | 0.79, 1.04 | 0.153 | 0.96 | 0.84, 1.09 | 0.520 | 1.03 | 0.90, 1.18 | 0.645 |
| Religious diversity index (RDI) | 0.85 | 0.72, 1.01 | 0.067 | 0.88 | 0.74, 1.04 | 0.132 | 0.87 | 0.74, 1.02 | 0.081 | 0.84* | 0.72, 0.99 | 0.039 | 0.86 | 0.73, 1.01 | 0.062 |
| Republican (%) | 1.32** | 1.12, 1.57 | 0.001 | 1.38*** | 1.17, 1.64 | <0.001 | 1.42*** | 1.20, 1.69 | <0.001 | 1.39*** | 1.17, 1.65 | <0.001 | 1.34** | 1.13, 1.60 | 0.001 |
| NME policies |  |  |  |  |  |  |  |  |  |  |  |  |  |  |  |
| (Base) | — | — |  | — | — |  | — | — |  | — | — |  | — | — |  |
| Relig NEM | 1.37 | 0.87, 2.14 | 0.168 | 1.25 | 0.80, 1.93 | 0.314 | 1.36 | 0.88, 2.08 | 0.160 | 1.15 | 0.75, 1.76 | 0.513 | 0.82 | 0.53, 1.25 | 0.348 |
| Relig + philo NME | 0.81 | 0.51, 1.28 | 0.376 | 0.74 | 0.47, 1.16 | 0.195 | 0.87 | 0.56, 1.36 | 0.550 | 0.76 | 0.49, 1.19 | 0.230 | 0.61* | 0.39, 0.95 | 0.029 |
| AUC | 0.73 |  |  | 0.72 |  |  | 0.72 |  |  | 0.72 |  |  | 0.72 |  |  |

<sup>†</sup> \*p<0.05; \*\*p<0.01; \*\*\*p<0.001
Abbreviations: CI = Confidence Interval, OR = Odds Ratio

#### Sensitivity Analysis of MMR Post-Pandemic Model Threshold

Our MMR Post-Pandemic Model examines factors associated with low county-level MMR rates, defined as rates below the commonly used 95% threshold, which is widely accepted as the minimum target for achieving herd immunity. ^17^ In Figures S7 and Table S7, we present sensitivity analyses for this threshold for the Post-Pandemic Model, to evaluate the robustness of our findings. Overall, our findings confirm the consistency and stability of the model outputs. However, we note that for the Post-Pandemic Model, the sensitivity analysis indicates that COVID-19 death rate is only significantly associated with high county-level MMR rates above the 95.0% threshold.

**Figure S7.**
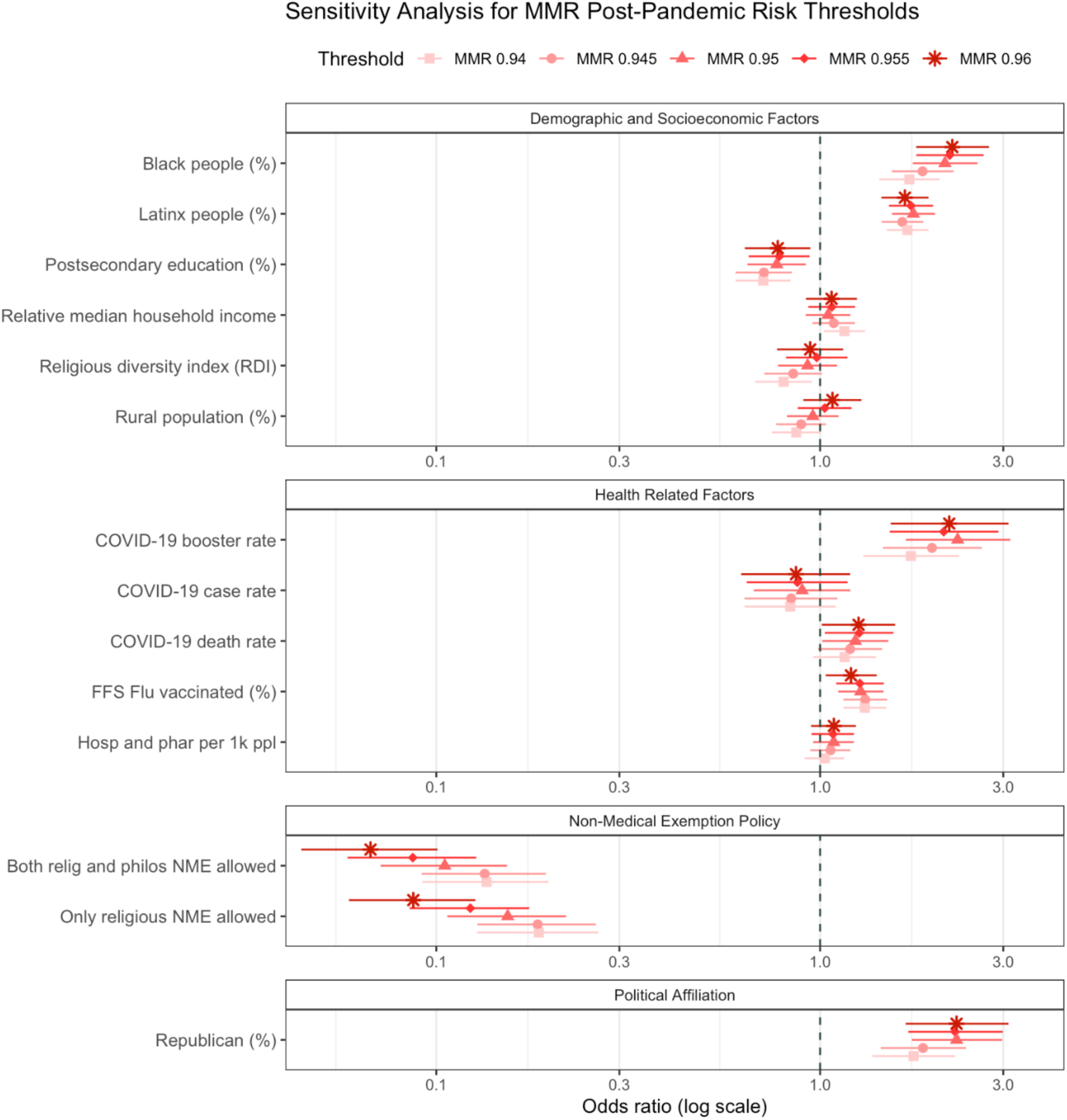
Results of the sensitivity analysis for the MMR Post-Pandemic Model. This figure illustrates the relationship between each explanatory factor and MMR vaccination rate as odds ratios, evaluating the following MMR thresholds: 94.0%, 94.5%, 95.0%, 95.5%, and 96.0%. The odds ratio of each threshold is represented by a corresponding shape in the legend, while the colored lines indicate the 95% confidence interval. An OR > 1 suggests that higher values of the factor are associated with counties maintaining MMR vaccination rates above the given threshold, whereas an OR < 1 suggests higher values are associated with counties having rates equal to or below the given threshold. The size of the baseline reference group for each threshold (i.e., the number of counties below each threshold) is as follows: 1323 (94.0%), 1414 (94.5%), 1515 (95%), 1619 (95.5%), 1709 (96.0%). Our findings confirm the general consistency and robustness of the results. However, we note that COVID-19 death rate is only significant above the 95.0% threshold.

**Table S7:** Presents the sensitivity analysis for the MMR Post-Pandemic Model, reporting odds ratios, 95% confidence intervals, and p-values for each explanatory factor. This table serves as a companion to Figure S7 and documents the relationship between each factor and MMR vaccination rates, evaluating the following MMR thresholds: 94.0%, 94.5%, 95.0%, 95.5%, and 96.0%. An OR > 1 suggests that higher values of the factor are associated with counties maintaining MMR vaccination rates above the given threshold, whereas an OR < 1 suggests higher values are associated with counties having rates below the given threshold. Statistical significance is denoted as follows: p <0.05 (*), p < 0.01 (**), p < 0.001 (***). The size of the baseline reference group for each threshold (i.e., the number of counties below each threshold) is as follows: 1323 (94.0%), 1414 (94.5%), 1515 (95%), 1619 (95.5%), 1709 (96.0%). Our findings confirm the consistency and robustness of the results.

| Characteristic | Post Model (0.94) |  |  | Model (0.945) |  |  | Model (0.95) |  |  | Model (0.955) |  |  | Model (0.96) |  |  |
| --- | --- | --- | --- | --- | --- | --- | --- | --- | --- | --- | --- | --- | --- | --- | --- |
|  | OR<sup>†</sup> | 95% CI | p-value | OR<sup>†</sup> | 95% CI | p-value | OR<sup>†</sup> | 95% CI | p-value | OR<sup>†</sup> | 95% CI | p-value | OR<sup>†</sup> | 95% CI | p-value |
| COVID-19 case rate | 0.84 | 0.64, 1.10 | 0.198 | 0.84 | 0.64, 1.11 | 0.220 | 0.90 | 0.67, 1.20 | 0.465 | 0.87 | 0.64, 1.18 | 0.377 | 0.87 | 0.62, 1.20 | 0.386 |
| COVID-19 death rate | 1.16 | 0.96, 1.40 | 0.126 | 1.20 | 0.99, 1.45 | 0.066 | 1.24* | 1.01, 1.51 | **0.036** | 1.26* | 1.03, 1.55 | **0.025** | 1.26* | 1.01, 1.57 | **0.039** |
| COVID-19 booster rate | 1.72*** | 1.30, 2.30 | **<0.001** | 1.96*** | 1.46, 2.64 | **<0.001** | 2.28*** | 1.67, 3.13 | **<0.001** | 2.10*** | 1.52, 2.92 | **<0.001** | 2.17*** | 1.53, 3.10 | **<0.001** |
| FFS Flu vaccinated (%) | 1.31*** | 1.15, 1.49 | **<0.001** | 1.31*** | 1.15, 1.50 | **<0.001** | 1.28*** | 1.11, 1.46 | **<0.001** | 1.27** | 1.10, 1.47 | **0.001** | 1.20* | 1.03, 1.40 | **0.018** |
| Hosp and phar per 1k ppl | 1.03 | 0.91, 1.16 | 0.650 | 1.06 | 0.94, 1.20 | 0.307 | 1.08 | 0.96, 1.22 | 0.189 | 1.08 | 0.95, 1.22 | 0.231 | 1.09 | 0.95, 1.24 | 0.227 |
| Black people (%) | 1.71*** | 1.43, 2.05 | **<0.001** | 1.85*** | 1.54, 2.23 | **<0.001** | 2.11*** | 1.74, 2.57 | **<0.001** | 2.18*** | 1.78, 2.67 | **<0.001** | 2.21*** | 1.78, 2.75 | **<0.001** |
| Latinx people (%) | 1.69*** | 1.49, 1.92 | **<0.001** | 1.64*** | 1.45, 1.86 | **<0.001** | 1.75*** | 1.54, 1.99 | **<0.001** | 1.72*** | 1.51, 1.97 | **<0.001** | 1.66*** | 1.44, 1.92 | **<0.001** |
| Postsecondary education (%) | 0.71*** | 0.60, 0.84 | **<0.001** | 0.71*** | 0.60, 0.84 | **<0.001** | 0.77** | 0.65, 0.92 | **0.004** | 0.78** | 0.65, 0.94 | **0.009** | 0.78* | 0.64, 0.94 | **0.011** |
| Relative median household income | 1.16* | 1.02, 1.31 | **0.021** | 1.08 | 0.95, 1.23 | 0.210 | 1.05 | 0.92, 1.20 | 0.482 | 1.07 | 0.93, 1.23 | 0.326 | 1.07 | 0.92, 1.25 | 0.377 |
| Rural population (%) | 0.87 | 0.75, 1.00 | 0.055 | 0.89 | 0.77, 1.04 | 0.136 | 0.96 | 0.82, 1.12 | 0.580 | 1.03 | 0.87, 1.21 | 0.738 | 1.08 | 0.90, 1.28 | 0.404 |
| Religious diversity index (RDI) | 0.80* | 0.68, 0.95 | **0.012** | 0.85 | 0.72, 1.01 | 0.066 | 0.93 | 0.78, 1.11 | 0.407 | 0.98 | 0.81, 1.18 | 0.830 | 0.94 | 0.77, 1.15 | 0.556 |
| Republican (%) | 1.75*** | 1.37, 2.25 | **<0.001** | 1.85*** | 1.44, 2.40 | **<0.001** | 2.27*** | 1.73, 2.98 | **<0.001** | 2.25*** | 1.70, 2.99 | **<0.001** | 2.27*** | 1.67, 3.10 | **<0.001** |
| NME policies |  |  |  |  |  |  |  |  |  |  |  |  |  |  |  |
| (Base) | — | — |  | — | — |  | — | — |  | — | — |  | — | — |  |
| Relig NEM | 0.18*** | 0.13, 0.26 | **<0.001** | 0.18*** | 0.13, 0.26 | **<0.001** | 0.15*** | 0.11, 0.22 | **<0.001** | 0.12*** | 0.09, 0.17 | **<0.001** | 0.09*** | 0.06, 0.13 | **<0.001** |
| Relig + philo NME | 0.14*** | 0.09, 0.20 | **<0.001** | 0.13*** | 0.09, 0.19 | **<0.001** | 0.11*** | 0.07, 0.15 | **<0.001** | 0.09*** | 0.06, 0.13 | **<0.001** | 0.07*** | 0.04, 0.10 | **<0.001** |
| AUC | 0.73 |  |  | 0.73 |  |  | 0.74 |  |  | 0.75 |  |  | 0.77 |  |  |

<sup>†</sup> \*p<0.05; \*\*p<0.01; \*\*\*p<0.001 Abbreviations: CI = Confidence Interval, OR = Odds Ratio

#### Sensitivity Analysis of Declining MMR Rate Thresholds

Figure S8 and Table S8 present the sensitivity analysis performed to evaluate the robustness of the -2.36% threshold used to define significant declines in MMR rates in the MMR Declining Model. Our findings confirm the general consistency and robustness of the results around the selected threshold. We observe that certain factors, such as rural population percentage, relative median household income, and FFS Flu vaccinated percent change in significance as the threshold varies. Specifically, rural population percentage becomes more significantly associated with declining MMR rates at larger declining thresholds, while relative median household income becomes more significantly associated with increasing MMR rates at larger declining thresholds. In contrast, FFS Flu Vaccinated percent loses significance at larger declining thresholds.

**Figure S8.**
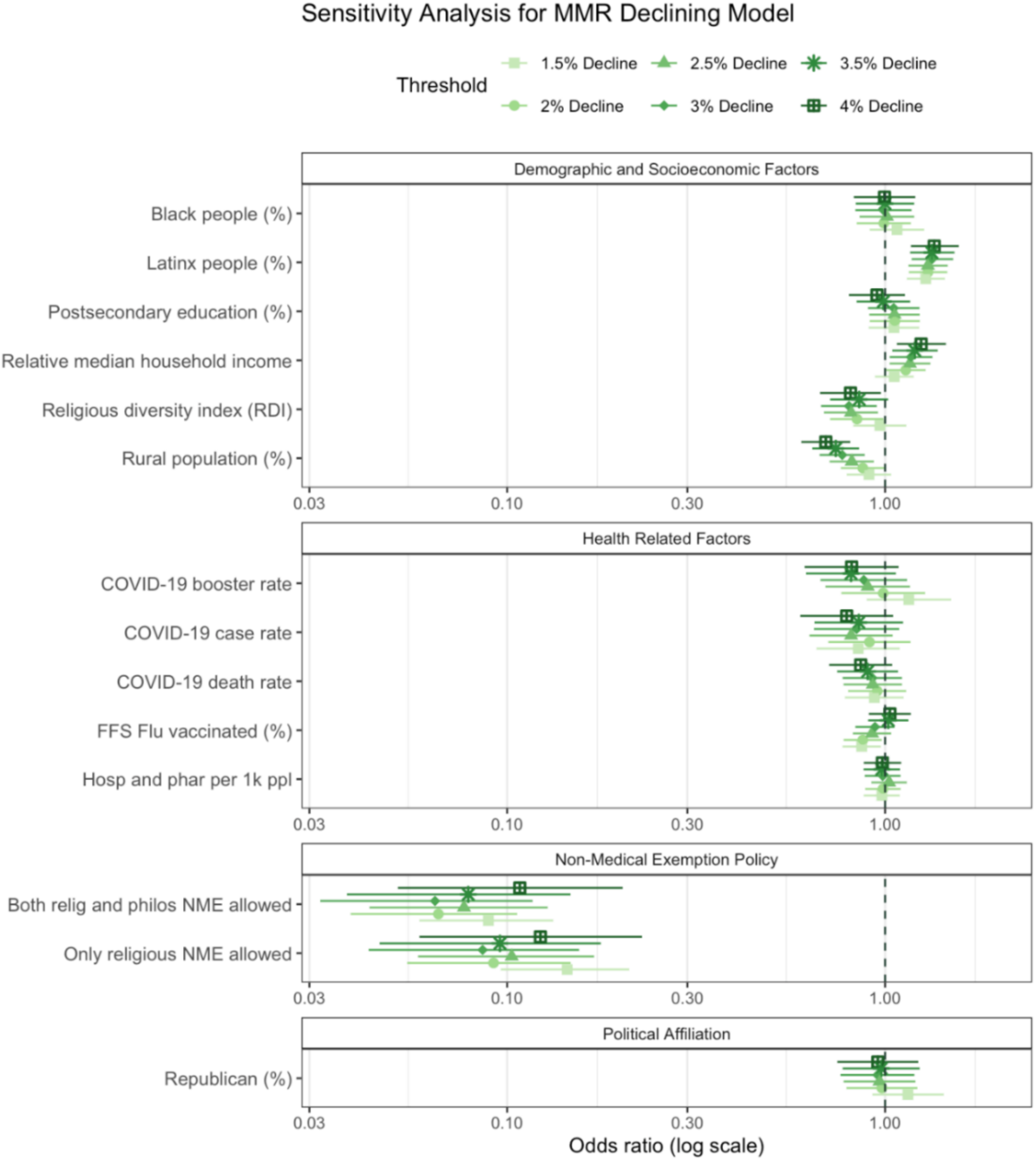
Results of the sensitivity analysis for the MMR Declining Model, which evaluates counties experiencing a substantial decline in MMR vaccination rates – defined as counties with declines greater than or equal to the national county-level average of -2.36%. This figure illustrates the relationship between each explanatory factor and declining MMR vaccination rates as odds ratios, evaluating thresholds representing the difference between pre- and post-pandemic MMR rates at: -1.5%, -2.0%, -2.5%, -3.0%, -3.5%, and -4.0%. Each threshold’s odds ratio is represented by a corresponding shape in the legend, while the colored lines indicate the 95% confidence interval. An OR > 1 suggests that counties with higher values of the factor are less likely to experience declining vaccination rates at the given threshold. Conversely, an OR < 1 suggests that counties with higher values of the factor are more likely to experience declining vaccination rates at the given threshold. The size of the baseline reference group for each threshold (i.e., the number of counties with declines ≥ the threshold) is as follows: 1251 (-1.5%), 1087 (-2.0%), 940 (-2.5%), 818 (-3.0%), 702 (-3.5%), and 602 (-4.0%).

**Table S8:** Presents the sensitivity analysis for the MMR Declining Model, which evaluates counties experiencing a significant decline in MMR vaccination rates – defined as counties with declines greater than the national county-level average of -2.36%. Serving as a companion to Figure S8, the table reports the relationship between each explanatory factor and MMR vaccination rates, evaluating thresholds representing the difference between pre- and post-pandemic MMR rates at: -1.5%, -2.0%, -2.5%, -3.0%, -3.5%, and -4.0%. For each threshold, the corresponding odds ratios, 95% confidence interval, and p-values are provided. An OR > 1 suggests that counties with higher values of the factor are less likely to experience declining vaccination rates at the given threshold. Conversely, an OR < 1 suggests that counties with higher values of the factor are more likely to experience declining vaccination rates at the given threshold. Statistical significance is denoted as follows: p <0.05 (*), p < 0.01 (**), p < 0.001 (***). The size of the baseline reference group for each threshold (i.e., the number of counties with declines ≥ the threshold) is as follows: 1251 (-1.5%), 1087 (-2.0%), 940 (-2.5%), 818 (-3.0%), 702 (-3.5%), and 602 (-4.0%).

| Characteristic | Decl Model (-1.5%) |  |  | Decl Model (-2.0%) |  |  | Decl Model (-2.5%) |  |  | Decl Model (-3.0%) |  |  | Decl Model (-3.5%) |  |  | Decl Model (-4.0%) |  |  |
| --- | --- | --- | --- | --- | --- | --- | --- | --- | --- | --- | --- | --- | --- | --- | --- | --- | --- | --- |
|  | OR<sup>1</sup> | 95% CI | p-value | OR<sup>1</sup> | 95% CI | p-value | OR<sup>1</sup> | 95% CI | p-value | OR<sup>1</sup> | 95% CI | p-value | OR<sup>1</sup> | 95% CI | p-value | OR<sup>1</sup> | 95% CI | p-value |
| COVID-19 case rate | 0.85 | 0.66, 1.09 | 0.205 | 0.91 | 0.71, 1.17 | 0.456 | 0.81 | 0.63, 1.05 | 0.109 | 0.84 | 0.65, 1.09 | 0.190 | 0.85 | 0.65, 1.12 | 0.244 | 0.79 | 0.60, 1.05 | 0.105 |
| COVID-19 death rate | 0.94 | 0.78, 1.12 | 0.473 | 0.95 | 0.80, 1.14 | 0.594 | 0.93 | 0.78, 1.11 | 0.408 | 0.93 | 0.77, 1.11 | 0.398 | 0.90 | 0.75, 1.08 | 0.262 | 0.86 | 0.71, 1.04 | 0.129 |
| COVID-19 booster rate | 1.16 | 0.89, 1.49 | 0.269 | 0.99 | 0.77, 1.28 | 0.929 | 0.90 | 0.70, 1.16 | 0.419 | 0.88 | 0.67, 1.14 | 0.334 | 0.81 | 0.62, 1.07 | 0.137 | 0.82 | 0.61, 1.09 | 0.162 |
| FFS Flu vaccinated (%) | 0.87* | 0.77, 0.97 | **0.016** | 0.87* | 0.78, 0.98 | **0.021** | 0.92 | 0.82, 1.04 | 0.182 | 0.94 | 0.83, 1.06 | 0.311 | 1.02 | 0.90, 1.15 | 0.756 | 1.03 | 0.91, 1.17 | 0.652 |
| Hosp and phar per 1k ppl | 0.98 | 0.88, 1.09 | 0.727 | 0.99 | 0.89, 1.10 | 0.815 | 1.02 | 0.92, 1.14 | 0.661 | 0.99 | 0.88, 1.10 | 0.787 | 0.98 | 0.88, 1.09 | 0.718 | 0.98 | 0.88, 1.10 | 0.776 |
| Black people (%) | 1.08 | 0.91, 1.27 | 0.390 | 0.99 | 0.84, 1.17 | 0.926 | 1.01 | 0.86, 1.19 | 0.898 | 0.99 | 0.83, 1.17 | 0.909 | 1.00 | 0.84, 1.19 | 0.994 | 1.00 | 0.83, 1.20 | 0.971 |
| Latinx people (%) | 1.28*** | 1.14, 1.44 | **<0.001** | 1.30*** | 1.16, 1.46 | **<0.001** | 1.30*** | 1.15, 1.46 | **<0.001** | 1.33*** | 1.18, 1.51 | **<0.001** | 1.33*** | 1.16, 1.53 | **<0.001** | 1.35*** | 1.17, 1.57 | **<0.001** |
| Postsecondary education (%) | 1.06 | 0.91, 1.23 | 0.489 | 1.06 | 0.91, 1.23 | 0.447 | 1.06 | 0.91, 1.23 | 0.463 | 1.05 | 0.90, 1.23 | 0.513 | 0.99 | 0.84, 1.17 | 0.903 | 0.95 | 0.80, 1.13 | 0.570 |
| Relative median household income | 1.06 | 0.94, 1.19 | 0.351 | 1.13* | 1.01, 1.28 | **0.040** | 1.16* | 1.03, 1.32 | **0.017** | 1.17* | 1.03, 1.33 | **0.018** | 1.20* | 1.04, 1.38 | **0.011** | 1.25** | 1.07, 1.45 | **0.004** |
| Rural population (%) | 0.91 | 0.79, 1.04 | 0.155 | 0.87* | 0.76, 1.00 | **0.046** | 0.82** | 0.71, 0.93 | **0.003** | 0.77*** | 0.67, 0.88 | **<0.001** | 0.74*** | 0.64, 0.85 | **<0.001** | 0.70*** | 0.60, 0.81 | **<0.001** |
| Religious diversity index (RDI) | 0.97 | 0.82, 1.14 | 0.698 | 0.84* | 0.72, 0.99 | **0.039** | 0.81* | 0.69, 0.96 | **0.014** | 0.80* | 0.68, 0.95 | **0.011** | 0.85 | 0.72, 1.02 | 0.080 | 0.81* | 0.67, 0.97 | **0.026** |
| Republican (%) | 1.15 | 0.93, 1.43 | 0.208 | 0.98 | 0.79, 1.22 | 0.859 | 0.97 | 0.78, 1.20 | 0.761 | 0.95 | 0.76, 1.20 | 0.686 | 0.98 | 0.77, 1.23 | 0.843 | 0.96 | 0.75, 1.22 | 0.725 |
| NME policies |  |  |  |  |  |  |  |  |  |  |  |  |  |  |  |  |  |  |
| (Base) | — | — | — | — | — | — | — | — | — | — | — | — | — | — | — | — | — | — |
| Relig NEM | 0.14*** | 0.10, 0.21 | **<0.001** | 0.09*** | 0.05, 0.15 | **<0.001** | 0.10*** | 0.06, 0.17 | **<0.001** | 0.09*** | 0.04, 0.16 | **<0.001** | 0.10*** | 0.05, 0.18 | **<0.001** | 0.12*** | 0.06, 0.23 | **<0.001** |
| Relig + philo NME | 0.09*** | 0.06, 0.13 | **<0.001** | 0.07*** | 0.04, 0.11 | **<0.001** | 0.08*** | 0.04, 0.13 | **<0.001** | 0.06*** | 0.03, 0.12 | **<0.001** | 0.08*** | 0.04, 0.15 | **<0.001** | 0.11*** | 0.05, 0.20 | **<0.001** |
| AUC | 0.65 |  |  | 0.66 |  |  | 0.66 |  |  | 0.68 |  |  | 0.68 |  |  | 0.69 |  |  |

<sup>1</sup> \*p<0.05; \*\*p<0.01; \*\*\*p<0.001
Abbreviations: CI = Confidence Interval, OR = Odds Ratio

#### Sensitivity Analysis of MMR Models Using Alternative Election Year Data

The U.S. elections data varies in 4-year cycles, as per the federal elections. To ensure that our findings regarding political affiliation were robust and not solely influenced by the election data used in our model (2020), we conducted a sensitivity analysis by substituting 2016 data for the Pre-Pandemic Model (Figure S9) and 2024 data for the Post-Pandemic Model (Figure S10). Our results remained consistent across both substitutions - with the exception of COVID-19 death rate in the post-pandemic model using 2024 data - reinforcing our finding that higher MMR rates are positively associated with Republican-aligned counties.

**Figure S9.**
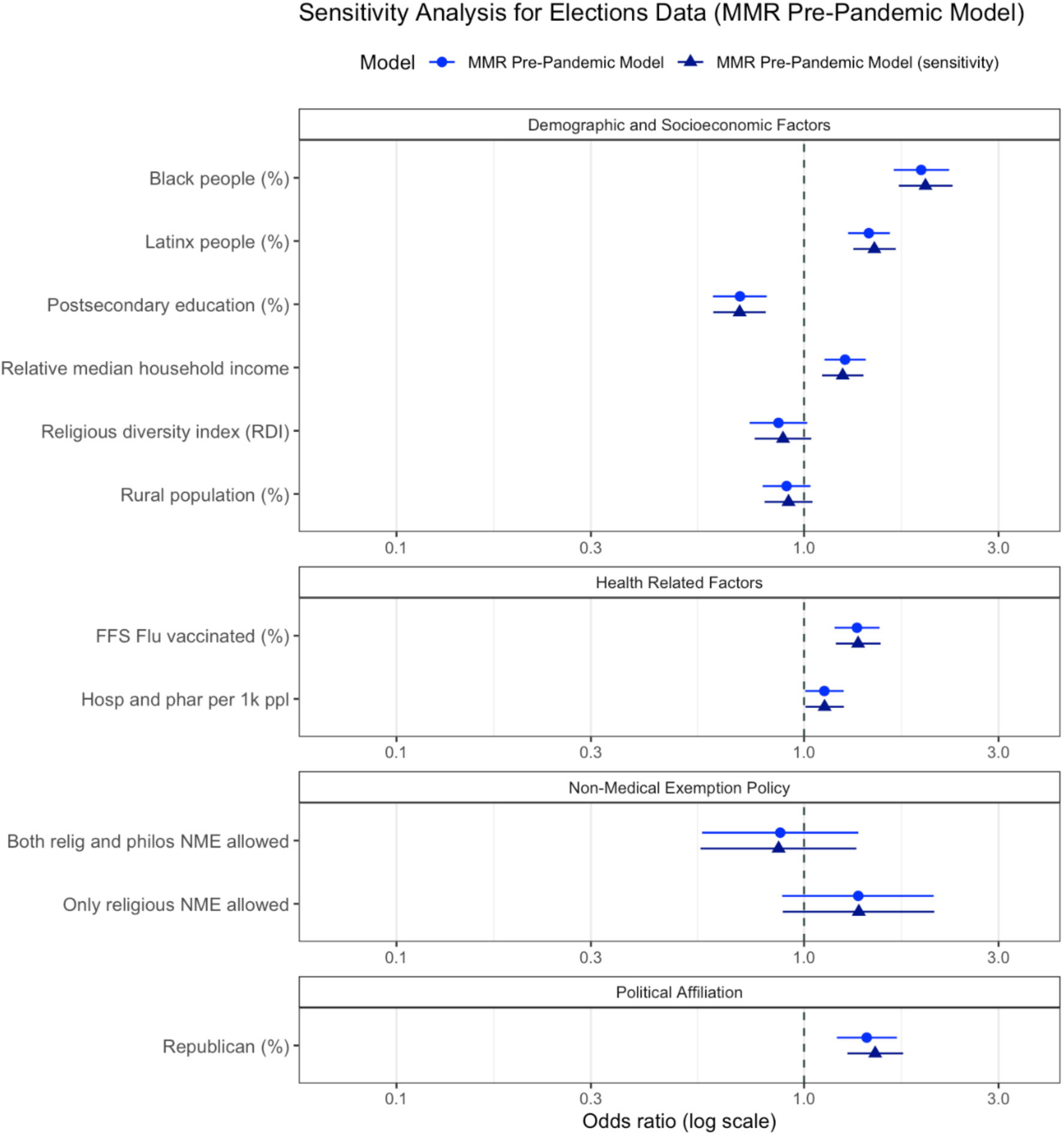
Results of the sensitivity analysis using 2016 and 2020 elections data in the Pre-Pandemic Model. This figure illustrates the relationship between each explanatory factor and MMR vaccination rate as odds ratios. The blue circle and line represent the odds ratios and corresponding 95% confidence interval for the original MMR Pre-Pandemic Model with 2020 elections data, whereas the blue triangle and line represent the odds ratios and corresponding 95% confidence interval for the MMR Pre-Pandemic Model where the 2020 election data has been replaced with the 2016 elections data. An OR > 1 suggests that higher values of the factor are associated with counties maintaining MMR vaccination rates above 95%, whereas an OR < 1 suggests higher values are associated with counties having rates below 95%.

**Figure S10.**
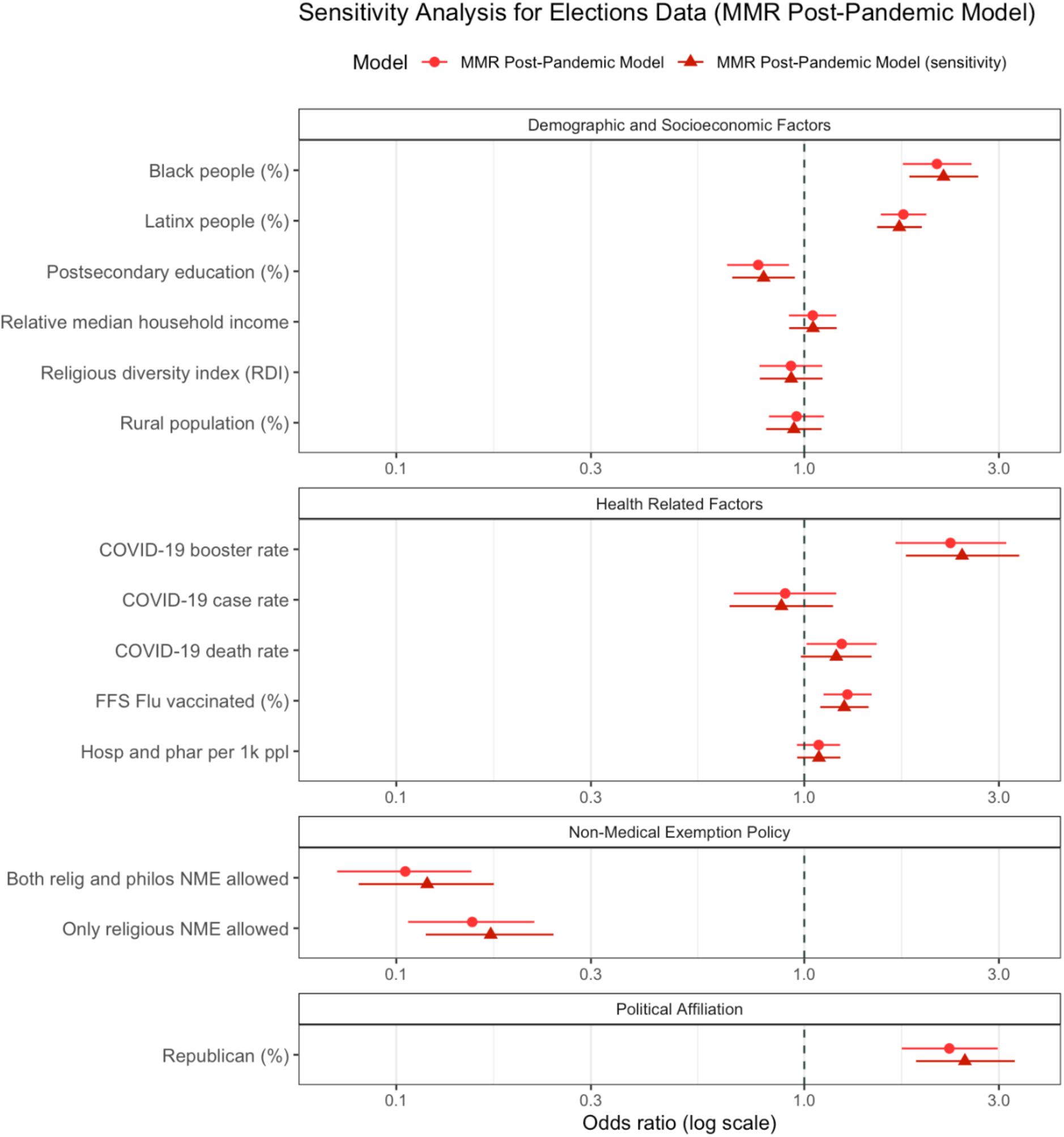
Results of the sensitivity analysis using 2020 and 2024 elections data in the Post-Pandemic Model. This figure illustrates the relationship between each explanatory factor and MMR vaccination rate as odds ratios. The red circle and line represent the odds ratios and corresponding 95% confidence interval for the original MMR Post-Pandemic Model with 2020 elections data, whereas the red triangle and line represent the odds ratios and corresponding 95% confidence interval for the MMR Post-Pandemic Model where the 2020 election data has been replaced with the 2024 elections data. An OR > 1 suggests that higher values of the factor are associated with counties maintaining MMR vaccination rates above 95%, whereas an OR < 1 suggests higher values are associated with counties having rates below 95%. With the exception of COVID-19 death rate, our findings remain consistent, reinforcing our conclusion that higher MMR rates are positively associated with Republican-aligned counties.

#### Joint Model Results

Table S9 documents the results of our combined logistic regression model. This model further reinforces our findings, as it reveals the same relationships observed in the independent MMR Pre-Pandemic and MMR Post-Pandemic Models. However, we chose to keep these models separate in our study, as we believe this approach better conveys the distinct factors associated with low MMR rates before and after the pandemic. We include the combined model in the supplementary materials to further illustrate the consistency and robustness of our results.

**Table S9.** Results for the combined logistic regression model, with time as an interaction term. The results of this model are consistent with the results of the Pre- and Post-Pandemic Models.

| Characteristic | OR<sup>†</sup> | 95% CI | p-value |
| --- | --- | --- | --- |
| FFS Flu vaccinated (%) | 1.35*** | 1.19, 1.53 | <0.001 |
| Hosp and phar per 1k ppl | 1.12* | 1.01, 1.25 | 0.040 |
| Black people (%) | 1.94*** | 1.66, 2.27 | <0.001 |
| Latinx people (%) | 1.44*** | 1.28, 1.63 | <0.001 |
| Postsecondary education (%) | 0.70*** | 0.60, 0.81 | <0.001 |
| Relative median household income | 1.26*** | 1.12, 1.42 | <0.001 |
| Rural population (%) | 0.91 | 0.79, 1.04 | 0.157 |
| Religious diversity index (RDI) | 0.87 | 0.73, 1.02 | 0.085 |
| Republican (%) | 1.42*** | 1.20, 1.69 | <0.001 |
| NME policies |  |  |  |
| (Base) | — | — |  |
| Relig NEM | 1.36 | 0.88, 2.09 | 0.164 |
| Relig + philo NME | 0.87 | 0.56, 1.36 | 0.554 |
| Period=post-covid | 1.10 | 0.57, 2.11 | 0.778 |
| (Period=post-covid) * FFS Flu vaccinated (%) | 0.95 | 0.79, 1.14 | 0.558 |
| (Period=post-covid) * Hospital accessibility | 0.97 | 0.82, 1.14 | 0.689 |
| (Period=post-covid) * Black people (%) | 1.09 | 0.85, 1.40 | 0.492 |
| (Period=post-covid) * Latinx people (%) | 1.21* | 1.02, 1.44 | 0.030 |
| (Period=post-covid) * Postsecondary education (%) | 1.11 | 0.88, 1.40 | 0.386 |
| (Period=post-covid) * Relative median household income | 0.83* | 0.70, 0.99 | 0.042 |
| (Period=post-covid) * Rural population (%) | 1.06 | 0.86, 1.30 | 0.600 |
| (Period=post-covid) * Religious diversity index (RDI) | 1.07 | 0.84, 1.36 | 0.570 |
| (Period=post-covid) * Republican (%) | 1.59** | 1.16, 2.19 | 0.004 |
| (Period=post-covid) * NME policies |  |  |  |
| (Period=post-covid) * Relig NEM | 0.11*** | 0.06, 0.20 | <0.001 |
| (Period=post-covid) * Relig + philo NME | 0.12*** | 0.07, 0.22 | <0.001 |
| (Period=post-covid) * COVID-19 case rate | 0.90 | 0.67, 1.19 | 0.461 |
| (Period=post-covid) * COVID-19 death rate | 1.24* | 1.02, 1.50 | 0.035 |
| (Period=post-covid) * COVID-19 booster rate | 2.28*** | 1.68, 3.12 | <0.001 |
| AUC | 0.76 |  |  |

<sup>†</sup> \*p<0.05; \*\*p<0.01; \*\*\*p<0.001 Abbreviations: CI = Confidence Interval, OR = Odds Ratio

---

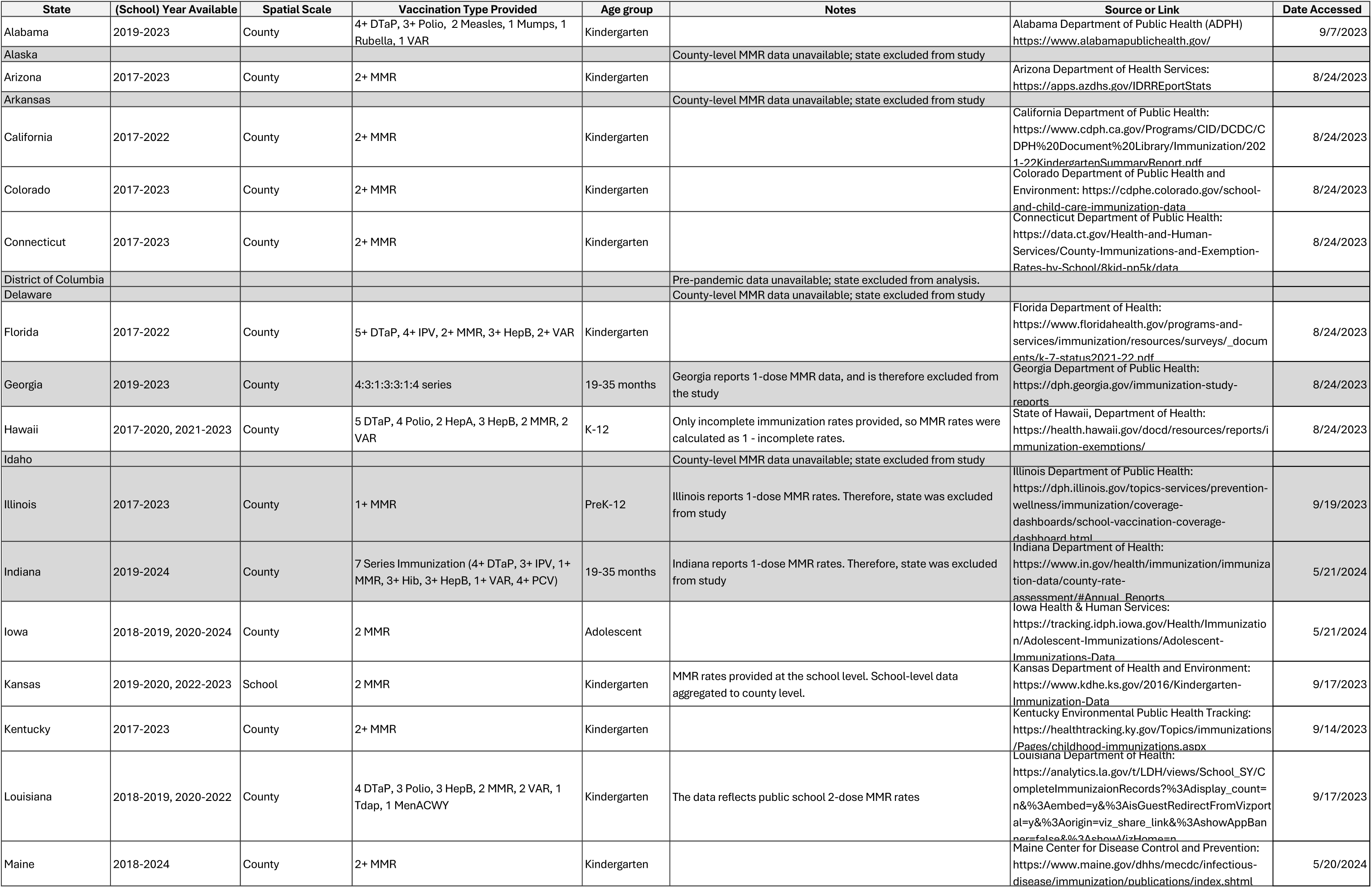
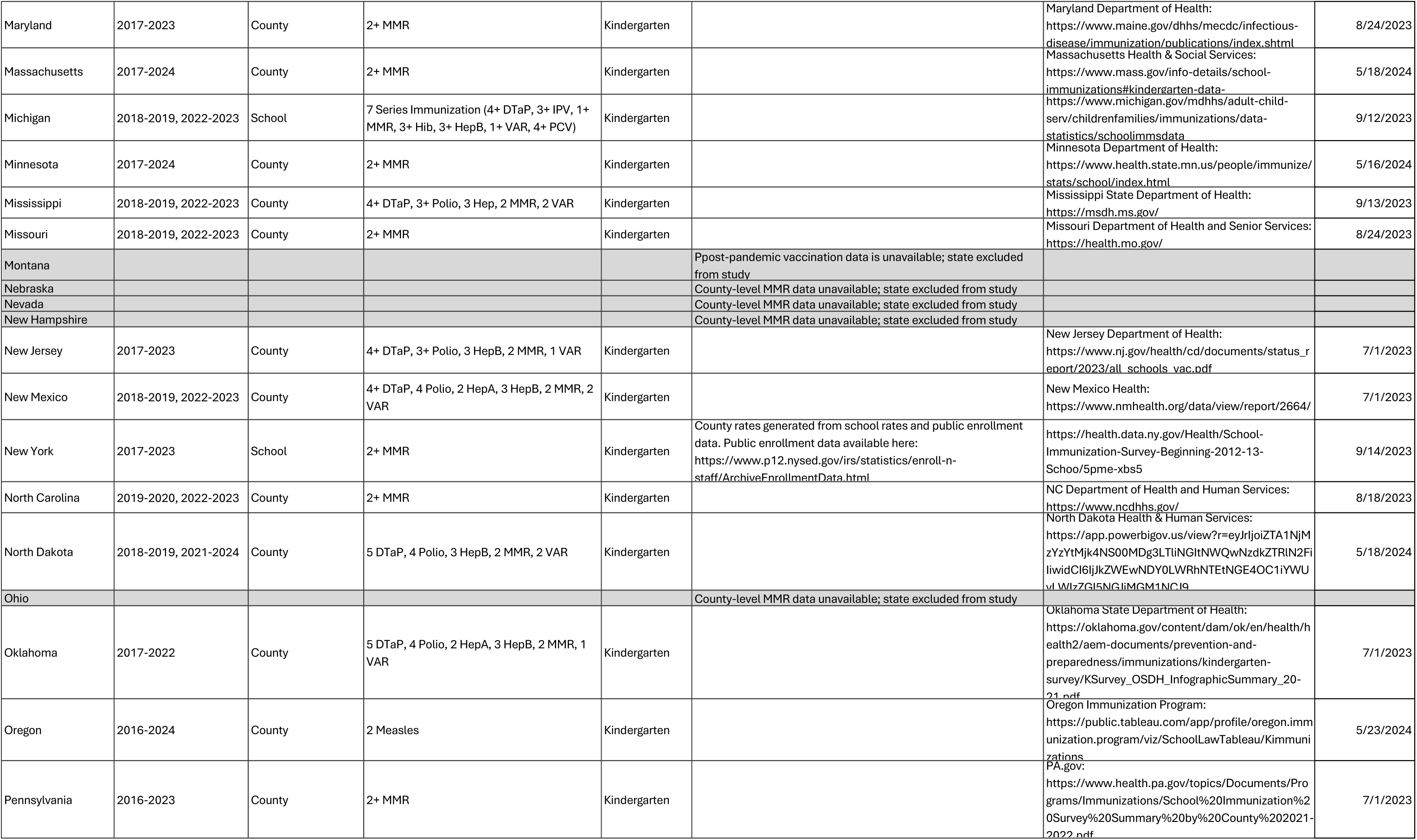
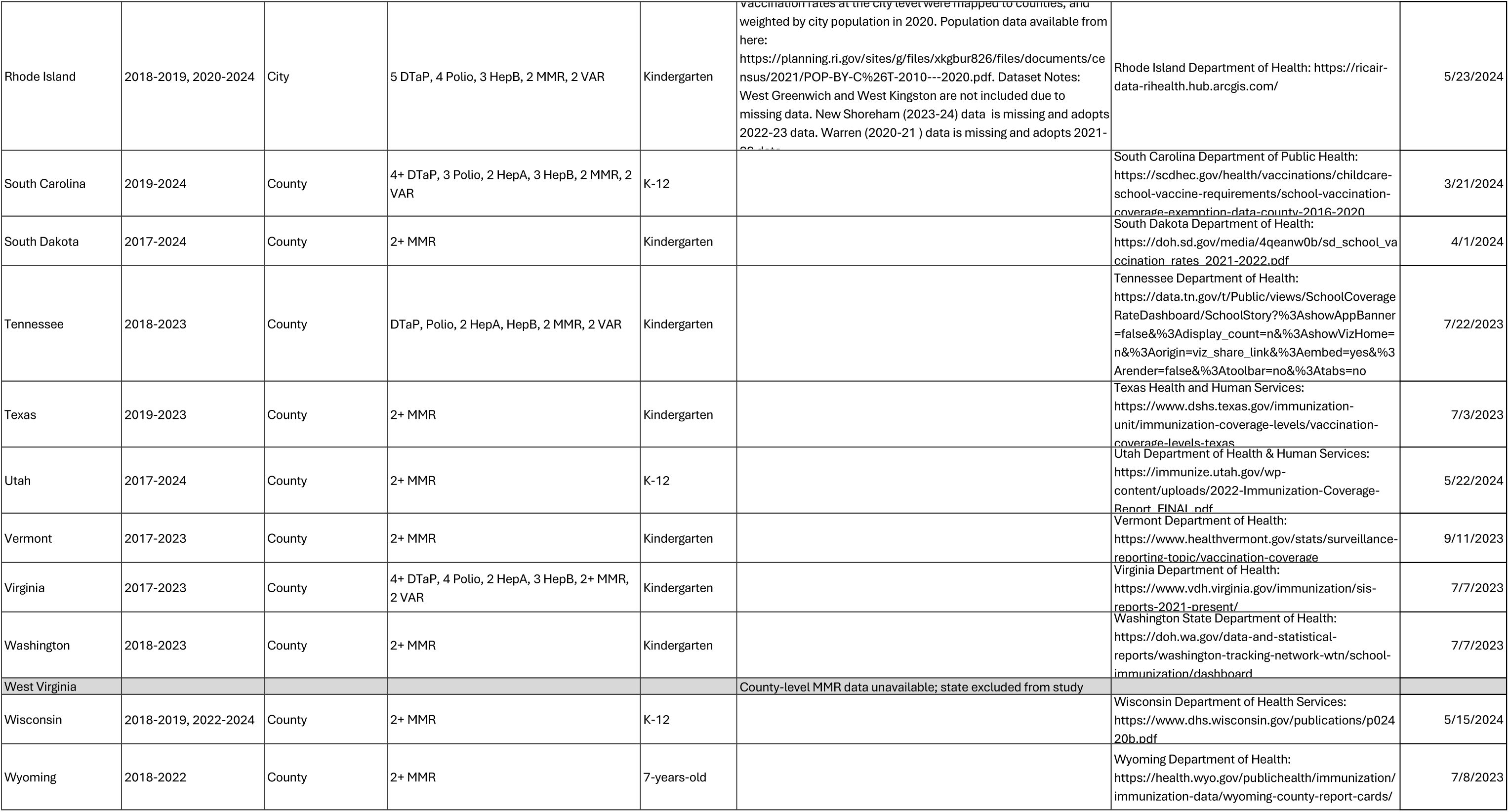

